# Does genetic liability for autism influence alcohol use?

**DOI:** 10.64898/2026.08.26.26360336

**Authors:** Stephanie Page, Kayleigh E Easey, Felicity Sedgewick, Dheeraj Rai, Evie Stergiakouli

## Abstract

A body of research suggests that autistic individuals are less likely to drink alcohol than neurotypicals. However, emerging studies support a link between autism and alcohol use. This complex relationship is also reflected in studies that have examined the genetic overlap between the two traits. However, it is unclear whether there is a direct causal relationship between them. To explore this, we applied a combination of polygenic score and Mendelian randomisation analyses using publicly available genome-wide summary statistics and phenotypic measures of autism and alcohol consumption from UK Biobank. LD score regression analyses did not provide evidence of a genetic correlation between genetic liability for autism and drinks consumed per week (r_g_=-0.08; CI95%=-0.19, 0.03). Further, findings from polygenic score analyses did not support an association between genetic liability for autism and overall monthly alcohol intake. Univariable Mendelian randomisation analyses showed little evidence for a total effect of autism, attention deficit hyperactivity disorder (ADHD) or depression on overall monthly alcohol consumption. Multivariable Mendelian randomisation analyses also showed little evidence of a direct effect of autism on drinks per week when controlling for ADHD and depression. It is plausible that genetic liability for autism does not directly increase the amount of alcohol consumed but instead operates via commonly co-occurring difficulties in the autistic community. However, our findings may be due to methodological shortcomings, including weak instruments biasing effects towards to the null. Consequently, results should be interpreted with caution and further research conducted to address these issues.

## 5.1 Introduction

Autism is a highly heritable form of neurodivergence in which individuals experience higher rates of physical and mental co-occurrences than the general population.^1,2^ One less-studied co-occurring issue in the autistic community with conflicting findings is alcohol use. Some suggest that compared to non-autistic individuals, autistic people are less likely to consume alcohol generally, or to drink in a way that poses rises to their health (i.e. hazardously), while others contradict this.^3–6^ Possible contributing factors include co-occurrences such as attention deficit hyperactivity (ADHD), depression and potential genetic overlap between autism and alcohol use (though findings for the latter are mixed).^7–11^ It is possible there is some overlap between autism and alcohol use genetically, but whether there is a direct causal effect of the former on the latter remains unclear.

To help us establish whether there is a direct causal effect of autism on alcohol use, we can draw upon multiple genetic epidemiological methods with differing sources of bias to strengthen inference.^12^ For example, Linkage disequilibrium score regression (LDSC) has been shown to robustly estimate genetic correlations (r_g_) between traits.^13–15^ However, genetic correlations cannot tell us anything about the direction of the relationship. Instead, we can estimate an individual’s genetic liability for a certain trait using Polygenic scores (PGS) and regress this on phenotypic outcomes. PGS denote an individual’s genetic liability to a specific trait that is calculated by summing the risk alleles identified in genome-wide association studies (GWAS) for that trait and weighting each by their corresponding effect sizes.^16^ Combining LDSC and PGS analyses when both have differing sources of bias can improve confidence in inferring a relationship between two traits.

Despite their advantages, neither LDSC or PGS can be used to infer causality. Instead, we can use Mendelian randomisation (MR), a form of instrumental variable analysis where genetic variants are used as proxy measures to estimate the total causal effect of an exposure on an outcome. MR can be used for causal inference by exploiting Mendel’s law of independent assortment whereby genetic variants are assorted independently of environment and other genetic factors (excluding those in linkage disequilibrium) during meiosis.^17^ MR is not hindered by reverse causation like observational methods because developed traits do not influence inherited genetic variants.^18^ Though MR is subject to other forms of potential bias (e.g. from weak instruments or horizontal pleiotropy) we can test for these when using it.^19^ This complements the use of PGS, where pleiotropic effects cannot be tested for due to the volume of variants included. Grouping MR with LDSC and PGS allows us to address the shortcomings of each while also triangulating observational evidence.

An extension of MR, multivariable MR (MVMR) can be used to isolate the direct effect of exposure on outcome while controlling for other exposures. ADHD and depression are two of the most common co-occurrences with autism.^1^ Observationally, both have been found to be associated with alcohol use in the population.^20,21^ Autistic individuals who are heavy drinkers have also reported higher depressive symptoms than lighter autistic drinkers, while those who have co-occurring ADHD are more likely to report AUD than those without ADHD.^3,4,22,23^ Therefore, it would be worthwhile to use MVMR to explore the direct causal effect of autism on alcohol use while controlling for ADHD and depression.

Clinically, and for the autistic community, it is important to know whether genetic liability for autism is a risk factor for drinking. It is also important to tease out the contributions of genetic liability for autism and commonly co-occurring conditions (such as ADHD and depression) on the risk for alcohol use. Further, it is essential to be able to test for pleiotropic effects that may influence effect estimates of genetic liability for autism on alcohol consumption. Given that alcohol use precedes AUD and there is increasing evidence to suggest the two are strongly genetically correlated (ranging from r_g_=0.86 to 1), this study will focus on alcohol use.^24^ Understanding the relationship between alcohol use and autism will enable informed and appropriate support for members of the autistic community, as existing services for neurotypical individuals may be inaccessible. The first aim of this study is to investigate the causal relationship of genetic liability for autism on alcohol consumption using LDSC, PGS and MR. The second is to explore the direction of effect using bidirectional MR, while the third is to examine the direct causal effect of autism on alcohol use on while controlling for two commonly co-occurring conditions (ADHD and depression) using MVMR.

## 5.2 Methods

Due to limited availability, all data used were based on individuals of European ancestry.

### Data sources

#### Autism genome-wide association study (GWAS)

Instruments for genetic liability for autism used in MR and MVMR were derived from summary statistics from a multi-cohort GWAS meta-analysis, including Integrative Psychiatric Research (iPYSCH) and the Psychiatric Genomics Consortium (PGC).^25^ The full summary statistics of this were also used in LDSC. N=13,076 cases of autism spectrum disorder were identified according to codes from version 10 of the International Classification of Diseases (ICD-10) in iPSYCH.^26^ N=5,305 ASD diagnoses were identified in iPSYCH using criteria from the ICD and/or Diagnostic and Statistical Manual of Mental Disorders (DSM-V), providing N=18,381 cases. N=27,969 controls were included without any psychiatric diagnoses recorded in iPSYCH (N=22,664) or autism diagnoses identified for the PGC (N=5,305).^27^ Five loci were identified at genome-wide significance (p=5x10^-8^). Details of ethical approved obtained by iPSYCH and PGC are specified in the Methods section of the paper.^25^

#### Attention deficit hyperactivity disorder (ADHD) GWAS

Instruments for genetic liability for ADHD used in MR and MVMR were derived from genome wide association meta-analysis summary statistics (N=70,593 across 28 individual cohorts).^28^ The full summary statistics of this were also used in LDSC. ADHD symptoms were measured through a symptom score based on 20 instruments derived from the Achenbach System of Empirically Based Assessment (ASEBA) and Strengths and Difficulties Questionnaire (SDQ).^29,30^ These were rated by parents, teachers and self-reports. N=38,691 cases of diagnosed ADHD were identified through record of clinical diagnosis and/or use of ADHD-specific medication (vs. N=186,843 controls). Details of ethical approval or informed consent was obtained for each cohort are available in van der Laan et al.^31^

#### Depression GWAS

Instruments for genetic liability for depression used in MR and MVMR were derived from summary statistics from a GWAS meta-analysis of Major Depressive Disorder/MDD by the PGC.^32^ The full summary statistics of this were also used in LDSC. N=3,887,532 individuals were included from 76 cohorts. N=525,197 cases of depression were identified within the European subsample (vs. N=3,362,335 controls). Depression was identified using a combination of clinical interviews including ICD and DSM criteria (4% of the sample), ICD codes from electronic health records (54%), questionnaires (14%) and self-reported diagnosis (27%). Details of ethical approval and informed consent obtained for each cohort are available in PGC (2025).^32^

#### Alcohol consumption GWAS

Instruments for genetic liability for alcohol consumption used in MR and MVMR were derived from summary statistics from a GWAS by Schumann et al.^33^ The full summary statistics of this were also used in LDSC. N=70,460 participants were included in the discovery GWAS (and approximately N=105,000 after replication). The authors derived continuous measure of grams per alcohol consumed per day, which they estimated from participant reported data of drinks per week and the type of drinks consumed. Non-drinkers were excluded from the primary analysis. Details of ethical approval and informed consent obtained for each cohort are available in the article.^33^

### Phenotypic measures

Phenotypic measures were derived from UK Biobank, a UK-based prospective cohort of around 500,000 people recruited between 2006 and 2010 between 40 and 69 years.^34^ Further details on the study aims, design, sample characteristics, quality control (QC) measures and ethical approval are available on the UK Biobank website (www.ukbiobank.ac.uk), in Collins et al. and the UK Biobank Ethics and Governance Framework.^35,36^ For the following phenotypic measures included, the sample size was N=501,936.

#### Confounders

Participant sex was obtained at recruitment, or self-reported. Age was recorded via the date of birth given at the assessment centre of the first instance between 2006-2010. Twenty genetic principal component scores were generated for all participants.

#### Outcomes

Autism was measured through two binary self-report measures of answering ‘Yes’ to whether they (N=408), or a first-degree blood relative (N=5,332), had ever received a diagnosis of ‘Autism, Asperger’s or autism spectrum disorder’. Alcohol consumption was recorded through six self-reported categorical measures of monthly alcohol intake, six continuous measures of weekly alcohol intake and overall sum of the latter (the details of which can be found in Supplementary Table S2).

### Statistical analysis

#### LDSC

Full summary statistics were used from data sources mentioned above.^25,31,32,37^ Using the munge.py function in Python, summary statistics were filtered to align with HapMap3 and strand-ambiguous SNPs excluded where allele orientation was indeterminable. Formatted datasets for autism, ADHD, depression and alcohol use were then fed into Python together with pre-computed European LD scores downloaded from 1000 Genomes, and genetic correlations (r_g_) were calculated using the ldsc.py function. Analyses were carried out using the programme RStudio (version 2025.09.1+401) and Python (version 2.7 accessed externally using system calls in RStudio).

#### Autism PGS

The base sample consisted of N=46,530 individuals with European ancestry (GWAS summary statistics from Grove et al.)^25^. The target sample included N=501,936 individuals with European ancestry (genetic data from UK Biobank). To ensure independence, SNPs were clumped within a window of 250 kilobase pairs (kb) of each other and a linkage disequilibrium (LD) threshold (r^2^) of 0.1, using a subset of 10,000 randomly selected individuals from the target sample. Using a range of p-value thresholds (given in Supplementary Table S1), eight PGS for were created for each individual using a weighted sum of risk alleles present in the target data (that had been identified as risk alleles in the base data).^38^ Multiple thresholds were used because autism is a complex trait and therefore likely to be polygenic, meaning that genome-wide significant SNPs – though robustly associated – may only explain a small proportion of phenotypic variance.^39^ Progressively relaxing the p value threshold increased the number of SNPs in each score (with the aim of increasing the phenotypic variance explained). Scores were calculated using PLINK version 1.9.0-b.7.7 in the Swiss Army Knife (SAK) tool version 5.1.0 in the UK Biobank Research Analysis Platform (UKB-RAP) on DNAnexus.^40,41^ Regression analyses were conducted using the python-R feature in the JupyterLab package on UKB-RAP.

#### Association between genetic liability for autism and phenotypic measures of alcohol consumption

To explore the associations between genetic liability for autism and alcohol consumption, a series of regression models were run in which PGS of varying p-value thresholds (see Supplementary Table S1) were regressed on a measure of overall monthly alcohol intake as the outcome. Age at assessment, sex and genetic principal components were all controlled for in each model.^42^ Analyses were run using the python-R feature in JupyterLab package on UKB-RAP.

#### Univariable Mendelian randomisation (UVMR)

Initially, we used a two-sample UVMR framework (including inverse-variance weighted/IVW, MR-Egger, Weighted median and Weighted mode methods) to estimate the total causal effect of autism on alcohol consumption. UVMR analyses were also carried out to explore the effect of attention deficit hyperactivity disorder (ADHD) and depression on alcohol consumption for comparison. UVMR relies on three core assumptions, which are detailed in Supplementary Methods S1.^43^ To check that genetic variants were associated with each of the exposures, we used an F statistic to gauge instrument strength (where a value that exceeded 10 was interpreted as good instrument strength).^44^

To examine whether these genetic variants affected the outcome through a pathway other than via the exposure (indicating potentially horizontally pleiotropic effects), we generated a series of plots (specifically single-SNP, funnel and forest). Single-SNP plots were assessed to whether the removal of specific SNPs would change the overall exposure-outcome relationship.^45^ Funnel plots were scanned for asymmetry (which would indicate horizontal pleiotropy; Bowden et al., 2015. Forest plots were used to explore the individual effects of each SNP.^46^ Additionally, scatter plots were generated to visualise effect estimates across UVMR methods for comparison (with similar gradients/direction of graph lines providing reassurance of similar estimates across methods). Cochran’s Q statistics were also calculated for each exposure to provide estimates of heterogeneity in SNP effect estimates on the outcome. Additionally, MR-Egger intercepts were generated to screen for horizontal pleiotropy (evidence of a non-zero intercept would indicate this).^45^

To screen for the presence of directional pleiotropy, we employed MR-Egger regression to test for a non-zero intercept term.^47^ To remove any potential SNPs that primarily associated with the outcome rather than any of the three exposures, Steiger filtering was also utilised. No SNPs were removed for genetic liability for alcohol consumption when ADHD or depression were the outcome, but four SNPs were removed when genetic liability for autism was the outcome (NSNPS= 41 down to 37).

#### Bidirectional UVMR

To establish the direction of effect, we also carried out bidirectional UVMR in which genetic liability for alcohol consumption was the exposure while autism, ADHD and depression were the outcomes. As with the forward UVMR, methods included IVW-MR, MR-Egger, Weighted median and Weighted mode. Further, a series of plots (single SNP/leave-one-out, funnel, forest and scatter) were generated for the same reasons above. To remove any potential SNPs that primarily associated with the outcome rather than any of the three exposures, Steiger filtering was also utilised. No SNPs were removed for genetic liability autism or ADHD through this process, but fourteen SNPs were removed for depression (NSNPS= 198 down to 184).

#### Multivariable Mendelian randomisation (MVMR)

To explore the direct causal effect of genetic liability for autism on units of alcohol consumed per week while accounting for ADHD and depression, we used MVMR-IVW analyses. To account for limited genome-wide hits for autism, the p-value threshold was relaxed to p=5x10^-7^ (while ADHD and depression were the standard p=5x10^-8^). To ensure exposure instruments were independent from each other, data were clumped with an LD (r^2^) of 0.001 and harmonised. MVMR analyses in which ADHD and depression were exposures were also conducted to assess the direct causal effect of each on alcohol consumption while controlling for autism.

As with UVMR, MVMR operates under three core assumptions These are an extension of UVMR assumptions, and as described by Sanderson et al.^48^:

1. The ‘relevance’ assumption: Instruments should be associated with each exposure of interest, independent of the other exposure(s)
2. The ‘exclusion restriction’ assumption: Instruments should be independent of the outcome other than via each of the exposures
3. The ‘independence/exchangeability’ assumption: Instruments must be independent of any confounders of the exposures and outcome

To assess the ‘relevance’ assumption, we calculated the conditional F statistic to test instrument strength (indicated by a threshold greater than 10)^48^. To improve instrument strength, pairwise MVMR was also conducted with genetic liability for autism and ADHD, autism and depression and ADHD depression. To test the ‘exclusion restriction’ assumption, we used Cochran’s Q to screen for heterogeneity, with a statistic greater than the number of SNPs indicating potential horizontal pleiotropy. Q-statistic minimisation was used to obtain more robust estimates for each of the exposures following evidence heterogeneity. MVMR-Egger analyses were also run due to being a pleiotropy-robust method.^49^ To remove any potential SNPs that primarily associated with the outcome rather than any of the three exposures, Steiger filtering was also utilised. Twelve SNPs were removed through this process (NSNPS= 202 down to 190).

MVMR results reflect the effect on the outcome (alcohol consumption) corresponding to one standard deviation increase in the exposure (autism, ADHD or depression when controlling for the other two exposures). All MR analyses in this paper were run using STROBE-MR guidelines (Supplementary Methods S2).^50^ UVMR and MVMR analyses were carried out using RStudio (version 2025.09.1+401), with UVMR analyses being conducted using the TwoSampleMR package (version 0.6.22) and MVMR analyses using the MVMR package (version 0.4.1).

## 5.3 Results

### Genetic correlation between autism and alcohol consumption

Results of LDSC analyses are presented in Table 1. There was little evidence of a weak negative genetic correlation between autism and alcohol consumption (r_g_=-0.08, CI95%=-0.19, 0.03). By comparison, we found some evidence to suggest a weak negative genetic correlation between ADHD and alcohol consumption (r_g_=-0.07, CI95%=-0.15, -0.00), but little evidence to suggest a weak negative genetic correlation between depression and alcohol consumption (r_g_=-0.04, CI95%=-0.10, 0.01). There was strong evidence of moderate positive genetic correlations between autism and ADHD (r_g_=0.47, CI95%=0.40, 0.50), autism and depression (r_g_=0.34, CI95%=0.29, 0.39), and ADHD and depression (r_g_=0.56, CI95%=0.53, 0.60).

**Table 1.**
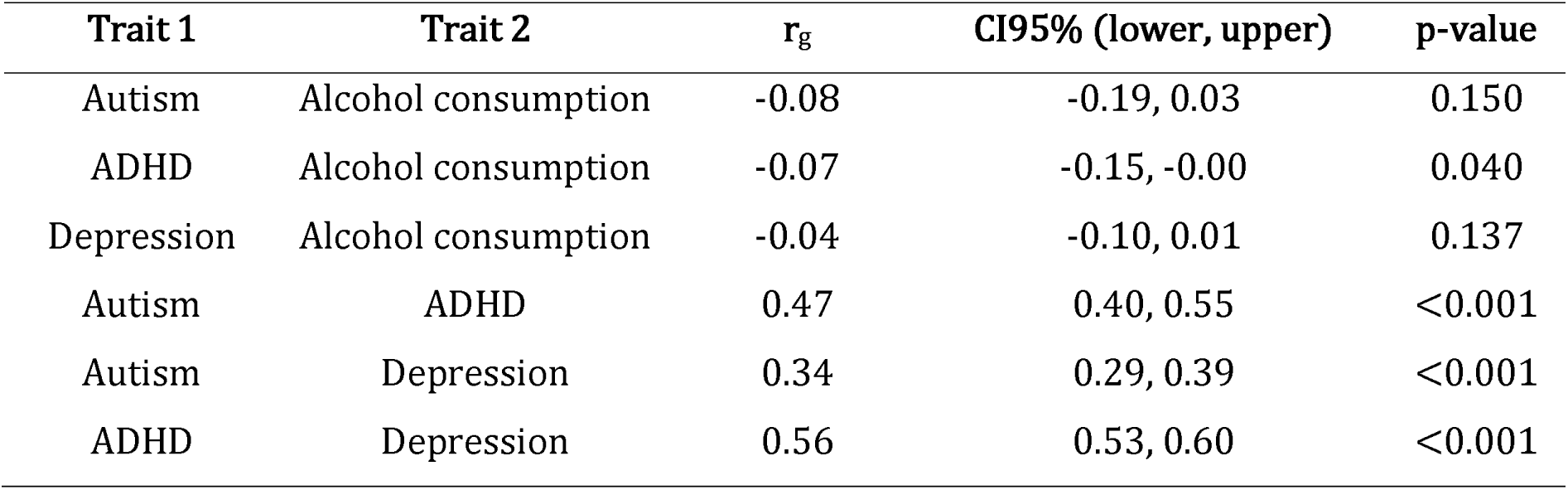
Results from LDSC estimating the genetic correlation (rg) between two traits.

| Trait 1 | Trait 2 | $r_g$ | CI95% (lower, upper) | p-value |
| --- | --- | --- | --- | --- |
| Autism | Alcohol consumption | -0.08 | -0.19, 0.03 | 0.150 |
| ADHD | Alcohol consumption | -0.07 | -0.15, -0.00 | 0.040 |
| Depression | Alcohol consumption | -0.04 | -0.10, 0.01 | 0.137 |
| Autism | ADHD | 0.47 | 0.40, 0.55 | <0.001 |
| Autism | Depression | 0.34 | 0.29, 0.39 | <0.001 |
| ADHD | Depression | 0.56 | 0.53, 0.60 | <0.001 |

**Table 2.** Results of linear regression analyses exploring the association between genetic liability for autism and overall monthly alcohol consumption measured in UK Biobank after controlling for multiple testing. Note: N=39,296 participants had data available for monthly alcohol consumption.

| Threshold | NSNPs | Beta | CI95% (lower, upper) | P value | R<sup>2</sup> | Adjusted R<sup>2</sup> | FDR q value |
| --- | --- | --- | --- | --- | --- | --- | --- |
| p1e-6 | 2 | -0.02 | -0.07, 0.02 | 0.284 | 0.0516 | 0.0510 | 0.512 |
| p1e-4 | 10 | -0.01 | -0.05, 0.04 | 0.779 | 0.0516 | 0.0510 | 0.876 |
| p1e-3 | 278 | -0.03 | -0.08, 0.01 | 0.174 | 0.0516 | 0.0510 | 0.392 |
| p0.01 | 10,562 | -0.04 | -0.10, 0.00 | 0.080 | 0.0516 | 0.0511 | 0.232 |
| p0.05 | 39,660 | -0.06 | -0.11, -0.02 | 0.010 | 0.0517 | 0.0512 | 0.088 |
| p0.1 | 70,922 | -0.05 | -0.09, 0.00 | 0.063 | 0.0516 | 0.0511 | 0.285 |
| p0.5 | 250,634 | -0.03 | -0.07, 0.02 | 0.309 | 0.0516 | 0.0510 | 0.512 |
| p1 | 381,610 | -0.03 | -0.07, 0.02 | 0.311 | 0.0516 | 0.0510 | 0.512 |

### Association between genetic liability for autism and overall monthly alcohol intake

Results of regression analyses exploring the associations between PGS reflecting genetic liability for autism and overall monthly alcohol intake are given in Figure 1. Linear regression analyses revealed evidence to suggest that genetic liability for autism (reflected in the “best-fit” PGS with the p-value threshold of 0.05) was associated with lower overall monthly alcohol intake (beta=-0.06, CI95%=-0.11,-0.02, p=0.010). Although there was no evidence of a relationship between any other PGS and overall monthly alcohol intake, beta values across thresholds were similar in effect size and direction (but confidence intervals were wider) (Figure 1).

**Figure 1.**
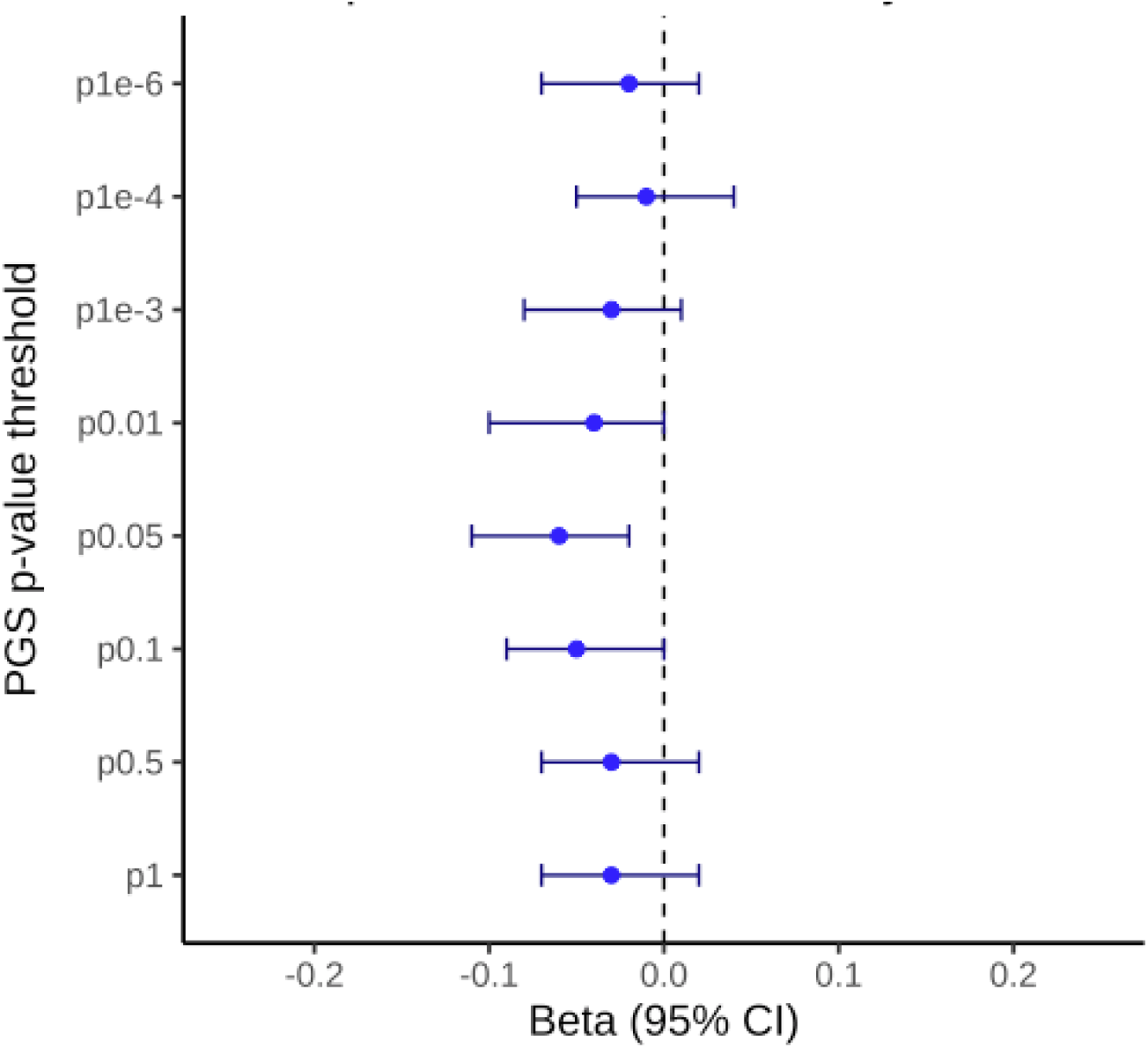
Forest plot of PGS effect estimates by p-value threshold after correcting for multiple testing.

### MR analyses

Results from univariable and multivariable Mendelian randomisation (UVMR and MVMR) analyses are reported in Tables 3 and 4.

**Table 3.**
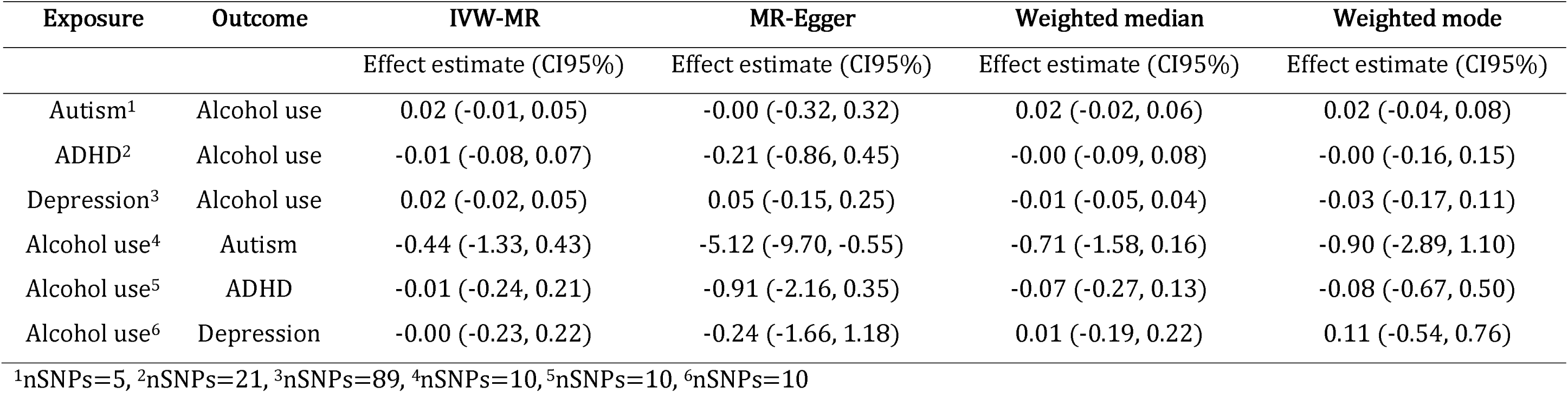
Results from bidirectional UVMR analyses of genetic liability for autism, attention deficit hyperactivity disorder (ADHD) and depression on units consumed per week (IVW-MR, MR-Egger, Weighted median and Weighted mode).

| Exposure | Outcome | IVW-MR | MR-Egger | Weighted median | Weighted mode |
| --- | --- | --- | --- | --- | --- |
|  |  | Effect estimate (CI95%) | Effect estimate (CI95%) | Effect estimate (CI95%) | Effect estimate (CI95%) |
| Autism<sup>1</sup> | Alcohol use | 0.02 (-0.01, 0.05) | -0.00 (-0.32, 0.32) | 0.02 (-0.02, 0.06) | 0.02 (-0.04, 0.08) |
| ADHD<sup>2</sup> | Alcohol use | -0.01 (-0.08, 0.07) | -0.21 (-0.86, 0.45) | -0.00 (-0.09, 0.08) | -0.00 (-0.16, 0.15) |
| Depression<sup>3</sup> | Alcohol use | 0.02 (-0.02, 0.05) | 0.05 (-0.15, 0.25) | -0.01 (-0.05, 0.04) | -0.03 (-0.17, 0.11) |
| Alcohol use<sup>4</sup> | Autism | -0.44 (-1.33, 0.43) | -5.12 (-9.70, -0.55) | -0.71 (-1.58, 0.16) | -0.90 (-2.89, 1.10) |
| Alcohol use<sup>5</sup> | ADHD | -0.01 (-0.24, 0.21) | -0.91 (-2.16, 0.35) | -0.07 (-0.27, 0.13) | -0.08 (-0.67, 0.50) |
| Alcohol use<sup>6</sup> | Depression | -0.00 (-0.23, 0.22) | -0.24 (-1.66, 1.18) | 0.01 (-0.19, 0.22) | 0.11 (-0.54, 0.76) |

<sup>1</sup>nSNPs=5, <sup>2</sup>nSNPs=21, <sup>3</sup>nSNPs=89, <sup>4</sup>nSNPs=10, <sup>5</sup>nSNPs=10, <sup>6</sup>nSNPs=10

**Table 4.** Results from IVW-MVMR analyses of genetic liability for autism, attention deficit hyperactivity disorder (ADHD) and depression on units consumed per week.

| Exposure | Outcome |  |  |  |
| --- | --- | --- | --- | --- |
|  |  | Effect estimate | CI95% (lower, upper) | P value |
| Autism | Alcohol use | -0.02 | -0.03, 0.06 | 0.231 |
| ADHD | Alcohol use | 0.06 | -0.04, 0.15 | 0.227 |
| Depression | Alcohol use | 0.01 | -0.03, 0.06 | 0.516 |

### Univariable MR (UVMR)

#### Total effects of genetic liability for autism on alcohol consumption

F statistics for genetic liability for autism, ADHD and depression were 28.10, 36.60 and 40.40, indicating good instrument strength (see Supplementary Table S3).^51^ Cochran’s Q statistics suggested some evidence of heterogeneity in SNP estimates for the ADHD and depression instruments (Supplementary Table S4). However, no evidence of non-zero intercepts was observed in any of the three exposure instruments tested through the MR Egger regression intercept term, providing reassurance against potential horizontal pleiotropy (see Supplementary Table S5).

As seen in Table 2, UVMR analyses revealed little evidence of a total effect of genetic liability for autism, ADHD or depression on alcohol consumption across any of the four methods (IVW-MR, MR-Egger, Weighted median or Weighted mode). Single SNP analyses revealed potential outliers for depression (rs10401087 and rs1160682; Supplementary Figure S9). No evidence of an effect was observed for depression following the removal of SNPs rs10401087 and rs1160682 (see Supplementary Tables S21-24). Funnel plots were symmetrical for autism, ADHD and depression (Supplementary Figures S2, S6 and S10). Forest plots for ADHD and depression (but not autism) showed SNPs with positive and negative effects, further indicating heterogeneity (Supplementary Figures S3, S7 and S11). Scatter plots provided reassurance of consistent effect estimates across methods (Supplementary Figures S4, S8 and S12).

### Bidirectional UVMR

#### Total effects of genetic liability of alcohol consumption on autism, ADHD and depression

The F statistic for genetic liability for alcohol consumption was 24.70, indicating good instrument strength (see Supplementary Table S3). There was some evidence of heterogeneity when genetic liability for alcohol consumption was the exposure for both ADHD and depression, but not autism, indicating potential horizontal pleiotropy (see Supplementary Table S4). Heterogeneity statistics did not differ after Steiger filtering (see Supplementary Tables S9).

As seen in Table 2, bidirectional UVMR analyses revealed little evidence of a total effect of genetic liability for alcohol consumption on autism, ADHD or depression across any of the four methods (IVW-MR, MR-Egger, Weighted median or Weighted mode). Single SNP analyses did not reveal any potential outliers for alcohol consumption as the exposure (Supplementary Figures S13, S17 and S21). Funnel plots were symmetrical where autism and ADHD were the outcome (Supplementary Figures S14 and S18), but asymmetrical for depression (Supplementary Figure S22). Forest plots with ADHD and depression as the outcome showed SNPs with both positive and negative effects, further indicating heterogeneity (Supplementary Figures S19 and S23). The forest plot where autism was the outcome showed evidence of a positive effect only (Supplementary Figure S15). A scatter plot where depression was the outcome provided reassurance of consistent effect across methods (Supplementary Table S24). Scatter plots where autism and ADHD were the outcome provided reassurance of consistent effect across IVW-MR, Weighted median and Weighted mode estimates, but not MR-Egger (Supplementary Figures S16 and S20). This could indicate potentially pleiotropic effects, however, there was little evidence of non-zero intercepts this in the MR-Egger intercept term (Supplementary Table S5). There was little evidence of any effects using bidirectional UVMR analyses after Steiger filtering (see Supplementary Tables S11-14).

### Multivariable MR (MVMR)

Conditional F statistics in MVMR analyses revealed scores below 10 for genetic liability for autism and ADHD, indicating poor instrument strength (2.83 and 6.65).^52^ Instrument strength appeared to be stronger for depression, with a conditional F statistic of 10.66 (see Supplementary Table S15). Cochran’s Q was greater than the number of SNPs (Q=115, p=0.050), indicating some evidence of heterogeneity (see Supplementary Table S16). Q-stat minimisation results (Qhet statistics) providing more robust effect estimates for each exposure are provided in Supplementary Table S17. Though the direction of effect for autism and depression instruments (but not ADHD) changed, the magnitude was very small for both (decrease of 0.03 for autism and an increase of 0.04 for depression). There was no evidence of an effect of any exposure on alcohol consumption observed after Steiger filtering (though effect direction changed for depression; see Supplementary Table 19). There was no evidence of an effect observed in any exposure in pairwise MVMR analyses (Supplementary Tables S27-S32).

Results of IVW-MVMR analyses in Table 3 show little evidence of a direct effect of genetic liability for autism on alcohol consumption when controlling for ADHD and depression (beta=-0.02, CI95%=-0.03, 0.06, p=0.231). MVMR-Egger analyses also showed little evidence of a direct effect of genetic liability for autism on alcohol consumption (beta=-0.02, CI95%=-0.04, 0.01, p=0.240) (Supplementary Table S18). IVW-MVMR analyses following Steiger filtering also showed little evidence of a direct effect of genetic liability for autism on alcohol consumption (beta=0.01, CI95%=-0.01, 0.03, p=0.538) (Supplementary Table S19). Steiger filtering marginally improved instrument strength for genetic liability for all three exposures (with an increase in conditional F statistic from 2.83 to 3.12 for autism, 6.65 to 7.36 for ADHD and 10.66 to 12.85 for depression; Supplementary Table S20). Evidence of heterogeneity (Cochran’s Q) disappeared (Q=36.80, p=0.999; Supplementary Table S21).

Results of IVW-MVMR analyses in Table 3 show little evidence of a direct effect of genetic liability for ADHD on alcohol consumption when controlling for autism and depression (beta=0.06, CI95%= -0.04, 0.15, p=0.227). There was also little evidence of a direct effect of genetic liability for depression on alcohol consumption when controlling for autism and ADHD (beta=0.01, CI95%= -0.03, 0.06, p=0.516).

## 5.4 Discussion

In this study we used a combination of LDSC, PGS, UVMR, bidirectional UVMR and MVMR analyses to explore the potential causal effect of genetic liability of autism on alcohol use. One polygenic score for genetic liability for autism was negatively associated with overall monthly alcohol intake, but the effect size was small. There was little evidence to suggest an effect of genetic liability for autism on monthly alcohol intake for any other polygenic score with differing p-value thresholds (though the direction of effects was the same across all, but confidence intervals were wider). These scores also explained similar levels of variance in alcohol consumption across all thresholds (which was also minimal). UVMR analyses showed little evidence for a total effect of genetic liability for autism, ADHD or depression on alcohol consumption and vice versa in bidirectional analyses. MVMR analyses showed little evidence of a direct effect of genetic liability for autism on alcohol consumption when controlling for ADHD and depression.

Observationally, there is conflicting evidence around the relationship between autism and alcohol use, with some studies supporting a lack of alcohol consumption in the autistic community.^5,6,53–59^ Conversely, other studies have found some members of the community at risk of alcohol use and/or alcohol-related problems, possibly due to risk factors including increasing age, being female and co-occurring difficulties such as ADHD and affective disorders.^3,4,60^ Further, Page et al. reported higher alcohol consumption in heavy drinkers with more social communication differences compared to those with less social communication differences from adolescence going into young adulthood.^61^ Together these observational findings suggest that being autistic is not necessarily a risk for alcohol use, but difficulties commonly associated with autism may increase this risk for some members in the autistic community. This may explain why we have not observed strong evidence of a total or direct causal effect of autism on alcohol consumption.

Genetically, there is some evidence to suggest shared genetic pathways between autism and alcohol consumption – though findings remain limited.^7,9–11,62^ Given that we observed evidence of heterogeneity in our MR analyses, it is possible that shared genetic pathways exist not just between autism and alcohol use alone, but with ADHD and depression too. However, despite attempts to address this (through methods including Steiger filtering, MR and MVMR-Egger, Qhet statistics and removal of SNPs potentially introducing noise), there still remained little evidence of a causal effect of any of the exposures of alcohol consumption (or vice versa). It is therefore difficult interpret what these pathways are based on the current findings.

### Strengths and limitations

#### Strengths

To the best of our knowledge, we used the largest publicly available genetic datasets for this type of analysis. F statistics were also greater than 10 for all three exposures in the univariable UVMR analyses, indicating good instrument strength.^44^ Further, although there was evidence of heterogeneity in the Cochran’s Q statistics for UVMR analyses, MR-Egger intercept analyses provided reassurance against horizontal pleiotropy (which would violate the “exclusion restriction” assumption). Further, the lack of evidence for a relationship between genetic liability for autism and alcohol use across results is consistent, strengthening inference. This also suggests that being genetically predisposed to autism does not influence an individual’s relationship with alcohol consumption, but instead modifiable non-genetic risk factors that lie on the pathway between them may be accountable. This would explain the variation in results of observational studies examining alcohol use in autistic individuals and is would be logical given the diversity of the autistic community. Clinically, this provides hope that there are opportunities for support to address any modifiable non-genetic risk factors.

#### Limitations

This study has several limitations. Firstly, as we can see in the LDSC results in Table 1, autism, ADHD and depression are all correlated genetically. This is problematic for UVMR, as one of the assumptions is that the instrument is not related to outcome via any pathway other than the exposure. UVMR results should therefore before interpreted with caution. Although MVMR analyses help address this by examining the direct effect of each exposure on the outcome, the MVMR analysis itself also carries limitations that may be the result of this. Cochran’s Q statistics were greater than the number of SNPs in MVMR analyses, indicating heterogeneity and therefore potentially pleiotropic effects. Further, conditional F statistics were less than 10 for two of the three exposures in MVMR analyses, indicating poor instrument strength that may bias effect estimates towards the null.^48^ Weak instruments can also evoke a false positive detection for heterogeneity due to pleiotropic effects when there is weak instrument bias.^48^ To address this, we used three approaches. Q-statistic minimisation was used to provide more robust IVW-MVMR estimates.^48^ MVMR-Egger analyses were also conducted to provide results from a pleiotropy-robust method for comparison.^63^ Additionally, Steiger filtering was applied to only include SNPs that were more strongly associated with each of the exposures than the outcome and both UVMR and MVMR analyses reran for comparison.^64^ None of these measures revealed evidence that contradicted our original findings, providing reassurance of robustness.

Further, given that UK Biobank is a general population study, the PGS calculated will have been comparatively underpowered than if a clinical autism study used. It is also possible that the self-reported phenotypic measures for alcohol consumption are subject to social desirability bias, or potential inaccuracies in quantities estimated by participants.^65^ Finally, the measures used in this study may be subject to selection bias. For example, the UK Biobank sample were more likely to be older, female and live in more affluent areas, as well as less likely to be obese, smoke or drink alcohol daily than individuals who did not participate.^66^ This sample is therefore not representative of the general population, limiting the generalisability of findings. Additionally, the genomic data used to calculate the polygenic scores only include individuals with European ancestry. Results therefore cannot be generalised to other populations due to the variation in genomic features between different ancestries.^67^

## Conclusion

In conclusion, our findings show little evidence of a total or direct causal effect of autism on alcohol consumption when controlling for ADHD and depression. This would suggest that any relationship between autism and alcohol intake is unlikely to be causal. However, it is also possible that our findings could be due to the identified methodical challenges. If this is the case, it is vital that further research is conducted to address the issues described. For example, future studies would benefit from drawing upon samples in which there is greater availability of phenotypic measures of autism, stronger instruments for autism and ADHD should they become available in future GWAS publications, and samples more representative of the general population. Addressing these may facilitate a clearer picture of how members of the autistic community are affected by this issue and what can be done to provide appropriate support.

## Data Availability

All data produced in the present study are available upon reasonable request to the authors

## Acknowledgements

The UK Biobank has approval from the North West Multi-centre Research Ethics Committee (REC reference: 16/NW/0274). This research was conducted using the UK Biobank resource under application number [110190]. All participants provided written informed consent.

## References

1. Khachadourian V, Mahjani B, Sandin S, et al. Comorbidities in autism spectrum disorder and their etiologies. Transl Psychiatry. 2023;13(1). doi:10.1038/S41398-023-02374-W

2. Zeidan J, Fombonne E, Scorah J, et al. Global prevalence of autism: A systematic review update. Autism Research. 2022;15(5):778–790. doi:10.1002/AUR.2696

3. Bowri M, Hull L, Allison C, et al. Demographic and psychological predictors of alcohol use and misuse in autistic adults. Autism. 2021;25(5):1469–1480. doi:10.1177/1362361321992668

4. Butwicka A, Långström N, Larsson H, et al. Increased Risk for Substance Use-Related Problems in Autism Spectrum Disorders: A Population-Based Cohort Study. J Autism Dev Disord. 2017;47(1):80–89. doi:10.1007/s10803-016-2914-2

5. Kaltenegger HC, Doering S, Gillberg C, Wennberg P, Lundström S. Low prevalence of risk drinking in adolescents and young adults with autism spectrum problems. Addictive Behaviors. 2021;113:106671. doi:10.1016/J.ADDBEH.2020.106671

6. Yule AM, DiSalvo M, Biederman J, et al. Decreased risk for substance use disorders in individuals with high-functioning autism spectrum disorder. Eur Child Adolesc Psychiatry. 2023;32(2):257–265. doi:10.1007/s00787-021-01852-0

7. Ahn Y, Kim J, Jung K, et al. Relationship Between Problematic Alcohol Use and Various Psychiatric Disorders: A Genetically Informed Study. 101176/appi.ajp20240095. 2025;182(7):671-682. doi:10.1176/APPI.AJP.20240095

8. Barber W, Aslan B, Meynen T, et al. Alcohol use among populations with autism spectrum disorder: narrative systematic review. BJPsych Open. 2025;11(1):e15. doi:10.1192/BJO.2024.824

9. Kranzler HR, Zhou H, Kember RL, et al. Genome-wide association study of alcohol consumption and use disorder in 274,424 individuals from multiple populations. Nature Communications 2019 10:1. 2019;10(1):1-11. doi:10.1038/s41467-019-09480-8

10. Mallard TT, Smoller JW. Beneath the Phenotypic Surface: Pleiotropy Shapes the Co-Occurrence of Alcohol Use Disorder and Psychopathology. 101176/appi.ajp20250411. 2025;182(7):593-595. doi:10.1176/APPI.AJP.20250411

11. Schumann G, Coin LJ, Lourdusamy A, et al. Genome-wide association and genetic functional studies identify autism susceptibility candidate 2 gene (AUTS2) in the regulation of alcohol consumption. Proc Natl Acad Sci U S A. 2011;108(17):7119–7124. doi:10.1073/PNAS.1017288108

12. Lawlor DA, Tilling K, Smith GD. Approaches to causal inference Triangulation in aetiological epidemiology. Int J Epidemiol. Published online 2016:1866–1886. doi:10.1093/ije/dyw314

13. Bulik-Sullivan B, Loh PR, Finucane HK, et al. LD Score regression distinguishes confounding from polygenicity in genome-wide association studies. Nat Genet. 2015;47(3):291–295. doi:10.1038/NG.3211

14. Chen J, Spracklen CN, Marenne G, et al. The trans-ancestral genomic architecture of glycemic traits. Nat Genet. 2021;53(6):840–860. doi:10.1038/S41588-021-00852-9

15. Lee SH, Yang J, Goddard ME, Visscher PM, Wray NR. Estimation of pleiotropy between complex diseases using single-nucleotide polymorphism-derived genomic relationships and restricted maximum likelihood. Bioinformatics. 2012;28(19):2540–2542. doi:10.1093/BIOINFORMATICS/BTS474

16. Garfield V, Anderson EL. A brief comparison of polygenic risk scores and Mendelian randomisation. BMC Med Genomics. 2024;17(1):10-. doi:10.1186/S12920-023-01769-4/FIGURES/1

17. Davey Smith G, Holmes M V., Davies NM, Ebrahim S. Mendel’s laws, Mendelian randomization and causal inference in observational data: substantive and nomenclatural issues. Eur J Epidemiol. 2020;35(2):99–111. doi:10.1007/S10654-020-00622-7

18. Mendelian Randomization - Book. Accessed August 12, 2026. https://www.mendelianrandomization.com/index.php/book-chapters

19. Hemani G, Tilling K, Davey Smith G. Orienting the causal relationship between imprecisely measured traits using GWAS summary data. PLoS Genet. 2017;13(11):e1007081. doi:10.1371/JOURNAL.PGEN.1007081

20. Luderer M, Ramos Quiroga JA, Faraone S V., Zhang James Y, Reif A. Alcohol use disorders and ADHD. Neurosci Biobehav Rev. 2021;128:648-660. doi:10.1016/J.NEUBIOREV.2021.07.010

21. Puddephatt JA, Irizar P, Jones A, Gage SH, Goodwin L. Associations of common mental disorder with alcohol use in the adult general population: a systematic review and meta-analysis. *Addiction (Abingdon*, England). 2022;117(6):1543–1572. doi:10.1111/ADD.15735

22. Huang JS, Yang FC, Chien WC, et al. Risk of Substance Use Disorder and Its Associations With Comorbidities and Psychotropic Agents in Patients With Autism. JAMA Pediatr. 2021;175(2):e205371–e205371. doi:10.1001/JAMAPEDIATRICS.2020.5371

23. Yule AM, DiSalvo M, Biederman J, et al. Decreased risk for substance use disorders in individuals with high-functioning autism spectrum disorder. Eur Child Adolesc Psychiatry. 2021;1:1–9. doi:10.1007/S00787-021-01852-0/TABLES/3

24. Johnson EC, Sanchez-Roige S, Acion L, et al. Polygenic contributions to alcohol use and alcohol use disorders across population-based and clinically ascertained samples. Psychol Med. 2020;51(7):1147. doi:10.1017/S0033291719004045

25. Grove J, Ripke S, Als TD, et al. Identification of common genetic risk variants for autism spectrum disorder. Nat Genet. 2019;51(3):431. doi:10.1038/S41588-019-0344-8

26. ICD-10 Version:2019. Accessed August 12, 2026. https://icd.who.int/browse10/2019/en

27. American Psychiatric Association. Diagnostic and Statistical Manual of Mental Disorders. Diagnostic and Statistical Manual of Mental Disorders. Published online May 22, 2013. doi:10.1176/APPI.BOOKS.9780890425596

28. van der Laan CM, Ip HF, Schipper M, et al. Publisher Correction: Genome-wide association meta-analysis of childhood ADHD symptoms and diagnosis identifies new loci and potential effector genes. Nat Genet. 2025;57(10):2604. doi:10.1038/S41588-025-02383-Z

29. Achenbach TM, Ruffle TM. The Child Behavior Checklist and related forms for assessing behavioral/emotional problems and competencies. Pediatr Rev. 2000;21(8):265–271. doi:10.1542/PIR.21-8-265

30. Goodman R. The Strengths and Difficulties Questionnaire: a research note. J Child Psychol Psychiatry. 1997;38(5):581–586. doi:10.1111/J.1469-7610.1997.TB01545.X

31. van der Laan CM, Ip HF, Schipper M, et al. Genome-wide association meta-analysis of childhood ADHD symptoms and diagnosis identifies new loci and potential effector genes. Nat Genet. 2025;57(10):2427–2435. doi:10.1038/s41588-025-02295-y

32. Adams MJ, Streit F, Meng X, et al. Trans-ancestry genome-wide study of depression identifies 697 associations implicating cell types and pharmacotherapies. Cell. 2025;188(3):640–652.e9. doi:10.1016/J.CELL.2024.12.002

33. Schumann G, Liu C, O’Reilly P, et al. KLB is associated with alcohol drinking, and its gene product β-Klotho is necessary for FGF21 regulation of alcohol preference. Proc Natl Acad Sci U S A. 2016;113(50):14372–14377. doi:10.1073/PNAS.1611243113

34. Allen NE, Sudlow C, Peakman T, Collins R. UK biobank data: Come and get it. Sci Transl Med. 2014;6(224). doi:10.1126/SCITRANSLMED.3008601/ASSET/A2997713-15D4-4A3A-865B-8B3FF0026AD8/ASSETS/GRAPHIC/6224ED4-FB.GIF

35. Collins R. What makes UK Biobank special? The Lancet. 2012;379(9822):1173–1174. doi:10.1016/S0140-6736(12)60404-8

36. UK BIOBANK Ethics and Governance Framework: Summary of comments on Version 1.0. Published online 2004.

37. Demontis D, Walters RK, Martin J, et al. Discovery of the first genome-wide significant risk loci for attention deficit/hyperactivity disorder. Nat Genet. 2019;51(1):63–75. doi:10.1038/S41588-018-0269-7

38. Choi SW, Mak TSH, O’Reilly PF. Tutorial: a guide to performing polygenic risk score analyses. Nature Protocols 2020 15:9. 2020;15(9):2759-2772. doi:10.1038/s41596-020-0353-1

39. Visscher PM, Wray NR, Zhang Q, et al. 10 Years of GWAS Discovery: Biology, Function, and Translation. Am J Hum Genet. 2017;101(1):5–22. doi:10.1016/J.AJHG.2017.06.005

40. Purcell S, Neale B, Todd-Brown K, et al. PLINK: a tool set for whole-genome association and population-based linkage analyses. Am J Hum Genet. 2007;81(3):559–575. doi:10.1086/519795

41. Research Analysis Platform - UK Biobank. Accessed August 12, 2026. https://www.ukbiobank.ac.uk/use-our-data/research-analysis-platform/

42. Choi SW, Mak TSH, O’Reilly PF. A guide to performing Polygenic Risk Score analyses. Nat Protoc. 2020;15(9):2759. doi:10.1038/S41596-020-0353-1

43. Davies NM, Holmes M V., Davey Smith G. Reading Mendelian randomisation studies: a guide, glossary, and checklist for clinicians. BMJ. 2018;362:601. doi:10.1136/BMJ.K601

44. Sanderson E, Davey Smith G, Windmeijer F, Bowden J. An examination of multivariable Mendelian randomization in the single-sample and two-sample summary data settings. Int J Epidemiol. 2019;48(3):713-727. doi:10.1093/ije/dyy262

45. Burgess S, Bowden J, Fall T, Ingelsson E, Thompson SG. Sensitivity Analyses for Robust Causal Inference from Mendelian Randomization Analyses with Multiple Genetic Variants. 30 | www.epidem.com Epidemiology •. 2017;28(1). doi:10.1097/eDe.0000000000000559

46. Rasooly D, Patel CJ. Conducting a Reproducible Mendelian Randomization Analysis Using the R Analytic Statistical Environment. Curr Protoc Hum Genet. 2019;101(1). doi:10.1002/CPHG.82

47. Bowden J, Smith GD, Burgess S. Mendelian randomization with invalid instruments: effect estimation and bias detection through Egger regression. Int J Epidemiol. 2015;44(2):512–525. doi:10.1093/IJE/DYV080

48. Sanderson E, Spiller W, Bowden J. Testing and correcting for weak and pleiotropic instruments in two-sample multivariable Mendelian randomization. Stat Med. 2021;40(25):5434–5452. doi:10.1002/SIM.9133

49. Grant AJ, Burgess S. Pleiotropy robust methods for multivariable Mendelian randomization. Stat Med. 2021;40(26):5813–5830. doi:10.1002/SIM.9156

50. Skrivankova VW, Richmond RC, Woolf BAR, et al. Strengthening the reporting of observational studies in epidemiology using mendelian randomisation (STROBE-MR): explanation and elaboration. BMJ. 2021;375. doi:10.1136/BMJ.N2233

51. Sanderson E, Glymour MM, Holmes M V., et al. Mendelian randomization. Nature reviews Methods primers. 2022;2(1):6. doi:10.1038/S43586-021-00092-5

52. Sanderson E, Smith GD, Windmeijer F, Bowden J. An examination of multivariable Mendelian randomization in the single-sample and two-sample summary data settings. doi:10.1093/ije/dyy262

53. Croen LA, Zerbo O, Qian Y, et al. The health status of adults on the autism spectrum. 101177/1362361315577517. 2015;19(7):814-823. doi:10.1177/1362361315577517

54. Fortuna RJ, Robinson L, Smith TH, et al. Health Conditions and Functional Status in Adults with Autism: A Cross-Sectional Evaluation. J Gen Intern Med. 2016;31(1):77–84. doi:10.1007/S11606-015-3509-X/TABLES/5

55. McLeod JD, Meanwell E, Hawbaker A. The Experiences of College Students on the Autism Spectrum: A Comparison to Their Neurotypical Peers. J Autism Dev Disord. 2019;49(6):2320–2336. doi:10.1007/S10803-019-03910-8/TABLES/5

56. Nylander L, Axmon A, Björne P, Ahlström G, Gillberg C. Older Adults with Autism Spectrum Disorders in Sweden: A Register Study of Diagnoses, Psychiatric Care Utilization and Psychotropic Medication of 601 Individuals. J Autism Dev Disord. 2018;48(9):3076-3085. doi:10.1007/s10803-018-3567-0

57. Russell AJ, Murphy CM, Wilson E, et al. The mental health of individuals referred for assessment of autism spectrum disorder in adulthood: A clinic report. 101177/1362361315604271. 2015;20(5):623-627. doi:10.1177/1362361315604271

58. Tolchard B, Stuhlmiller C. Chronic health and lifestyle problems for people diagnosed with autism in a student-led clinic. Advances in Autism. 2018;4(2):66–72. doi:10.1108/AIA-01-2018-0002/FULL/XML

59. Vohra R, Suresh Madhavan •, Sambamoorthi U. Emergency Department Use Among Adults with Autism Spectrum Disorders (ASD). J Autism Dev Disord. 2016;46. doi:10.1007/s10803-015-2692-2

60. Barber C. Meeting the healthcare needs of adults on the autism spectrum. Published online 2017.

61. Page S, Easey KE, Sedgewick F, Rai D, Stergiakouli E, Parker RMA. Autistic traits and alcohol consumption through adolescence and young adulthood. medRxiv. Published online November 25, 2025:2025.11.24.25340869. doi:10.1101/2025.11.24.25340869

62. Zuo L, Wang K, Zhang XY, et al. Association between common alcohol dehydrogenase gene (ADH) variants and schizophrenia and autism. Hum Genet. 2013;132(7):735. doi:10.1007/S00439-013-1277-4

63. Rees JMB, Wood AM, Burgess S. Extending the MR-Egger method for multivariable Mendelian randomization to correct for both measured and unmeasured pleiotropy. Stat Med. 2017;36(29):4705. doi:10.1002/SIM.7492

64. Hemani G, Tilling K, Davey Smith G. Orienting the causal relationship between imprecisely measured traits using GWAS summary data. PLoS Genet. 2017;13(11):e1007081. doi:10.1371/JOURNAL.PGEN.1007081

65. Davis CG, Thake J, Vilhena N. Social desirability biases in self-reported alcohol consumption and harms. Addictive behaviors. 2010;35(4):302–311. doi:10.1016/J.ADDBEH.2009.11.001

66. Fry A, Littlejohns TJ, Sudlow C, et al. Comparison of Sociodemographic and Health-Related Characteristics of UK Biobank Participants With Those of the General Population. Am J Epidemiol. 2017;186(9):1026–1034. doi:10.1093/AJE/KWX246

67. Moreno-Grau S, Vernekar M, Lopez-Pineda A, et al. Polygenic risk score portability for common diseases across genetically diverse populations. Hum Genomics. 2024;18(1). doi:10.1186/S40246-024-00664-Y

