## Supplementary Figures for "Does genetic liability for autism influence alcohol use?"

**The following figures represent analysis results prior to Steiger filtering.**

Supplementary Figure 1. Single SNP analysis from UVMR of autism on alcohol use

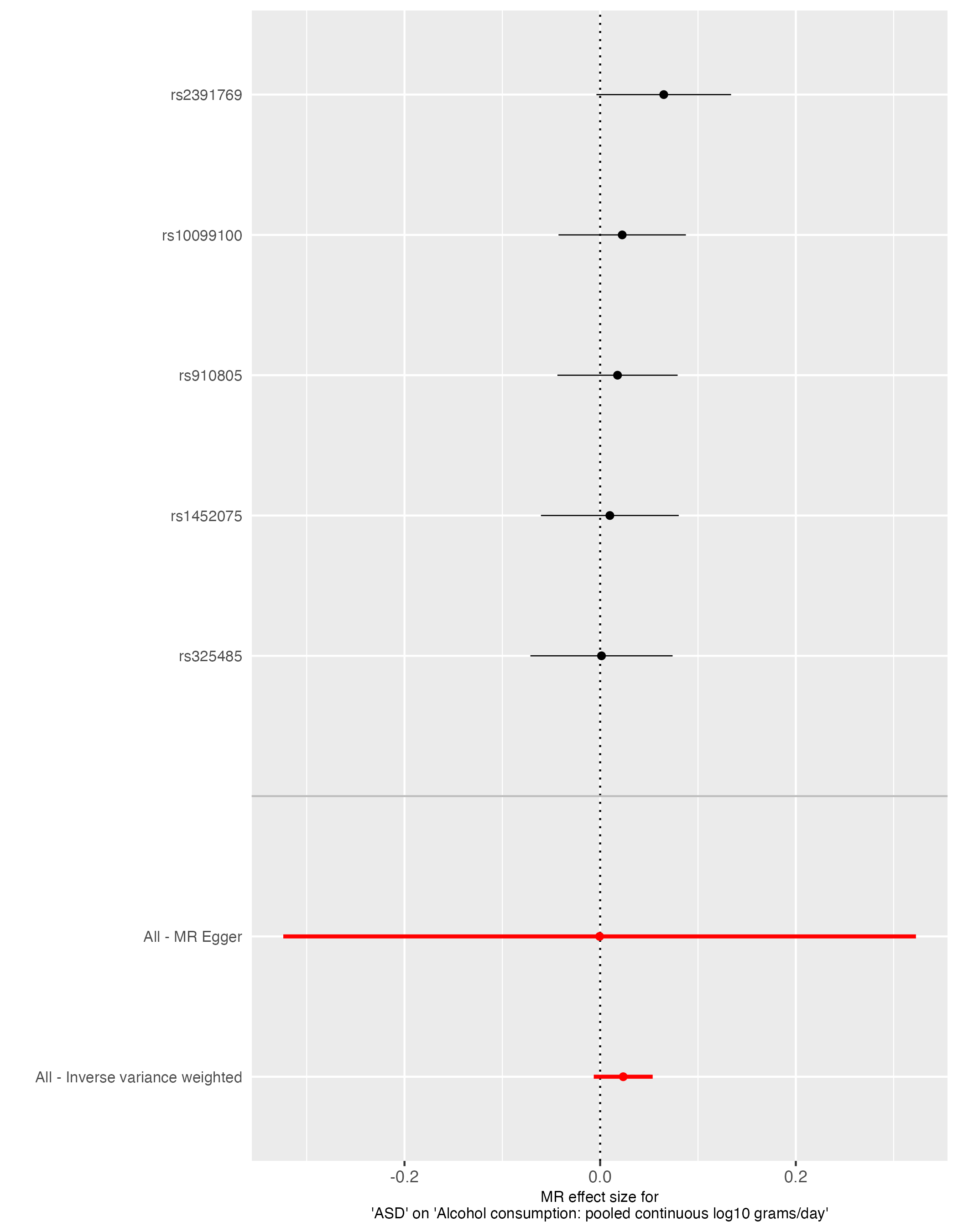

Supplementary Figure 2. Funnel plot from UVMR of autism on alcohol use

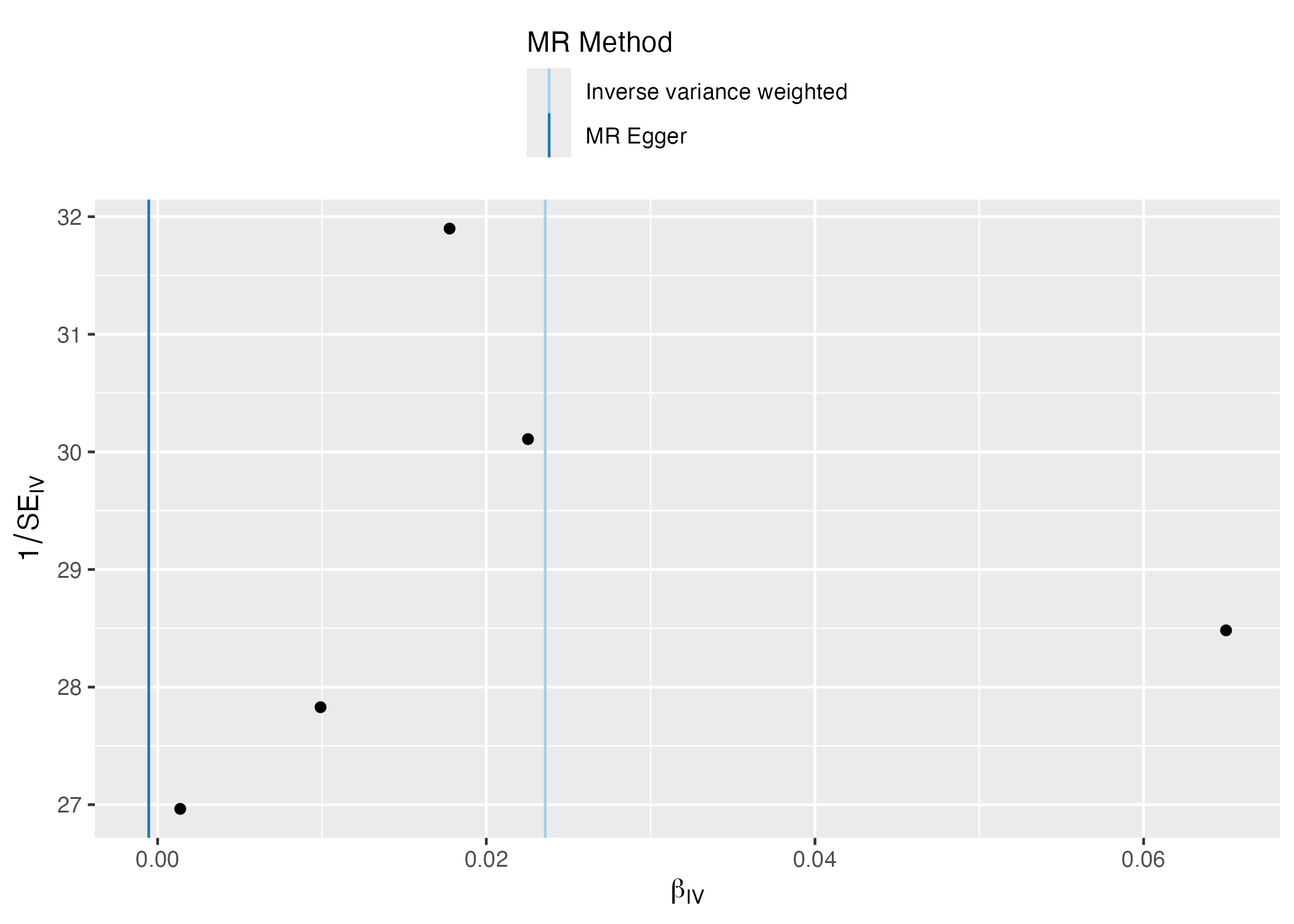

Supplementary Figure 3. Forest plot analysis from UVMR of autism on alcohol use

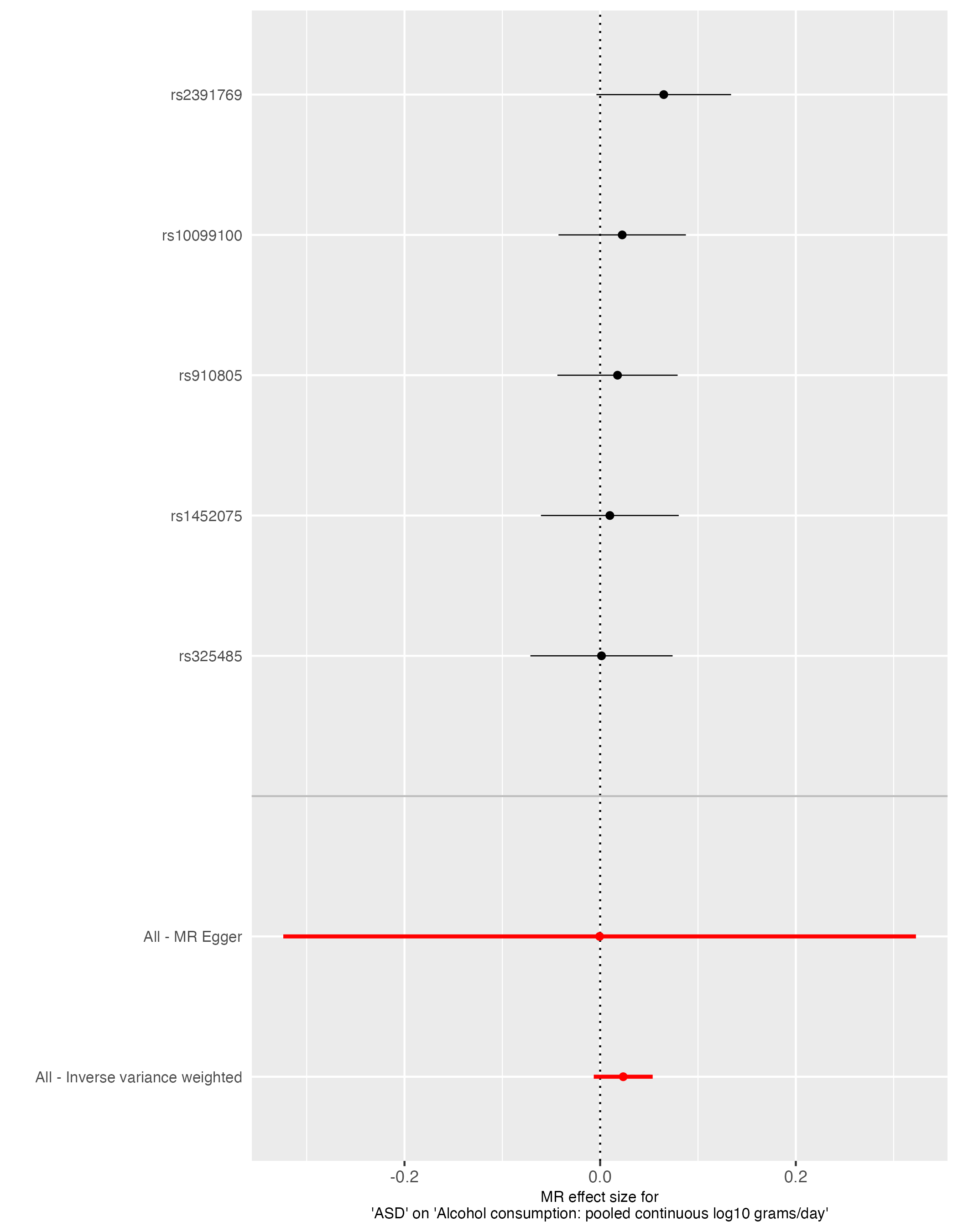

Supplementary Figure 4. Scatter plot analysis from UVMR of autism on alcohol use

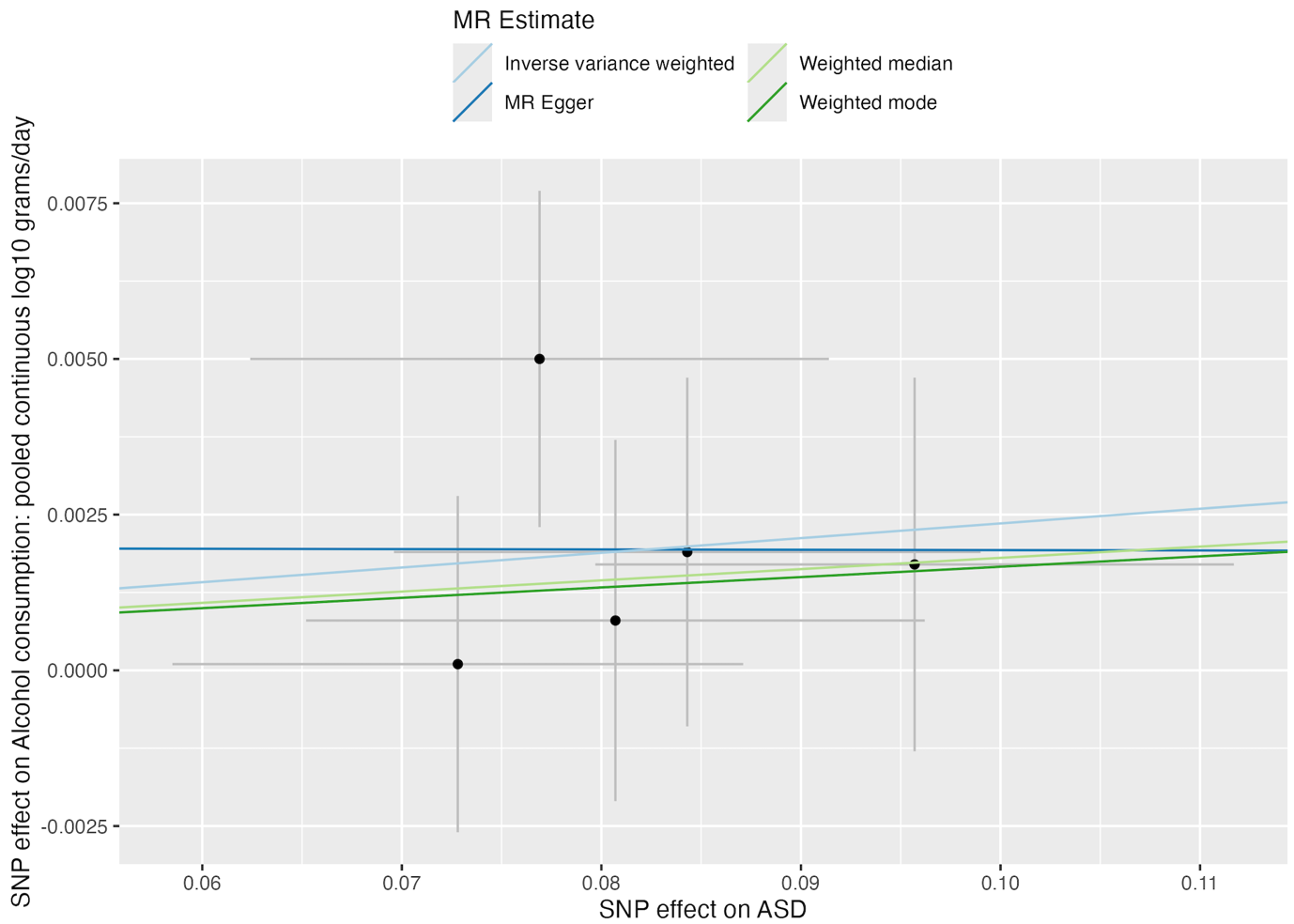

Supplementary Figure 5. Single SNP analysis from UVMR of attention deficit hyperactivity disorder (ADHD) on alcohol use

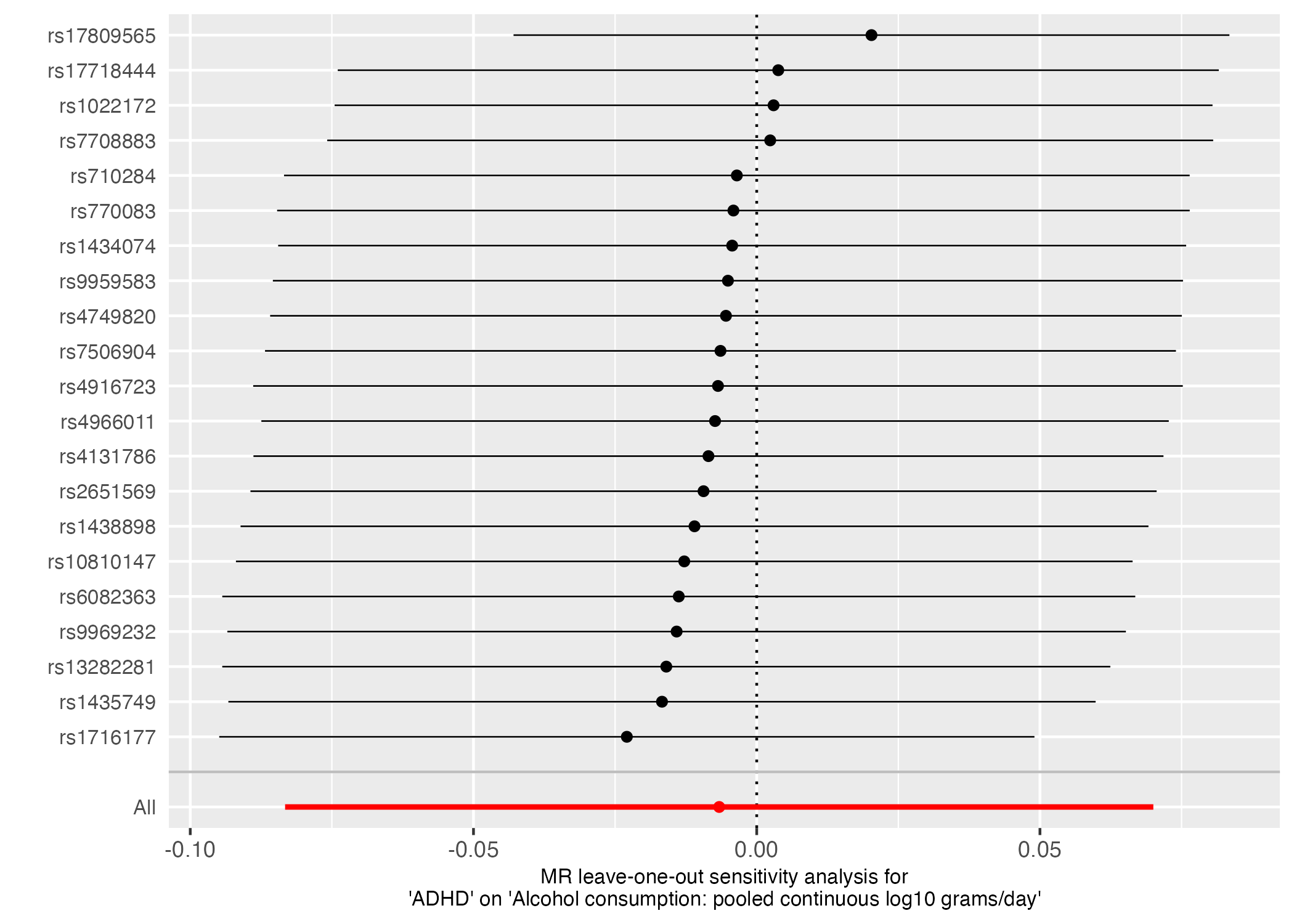

Supplementary Figure 6. Funnel plot from UVMR of attention deficit hyperactivity disorder (ADHD) on alcohol use

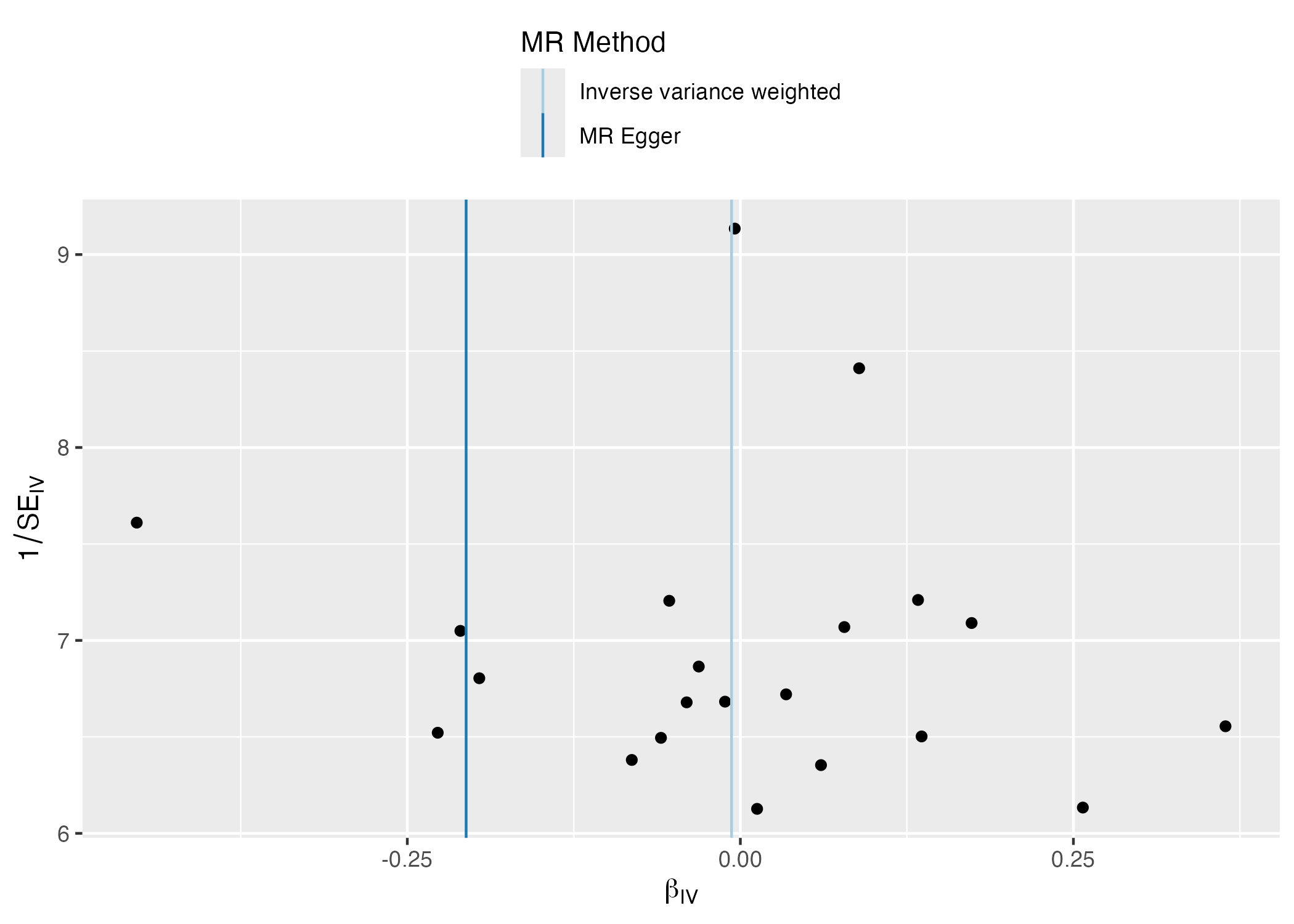

Supplementary Figure 7. Forest plot analysis from UVMR of attention deficit hyperactivity disorder (ADHD) on alcohol use

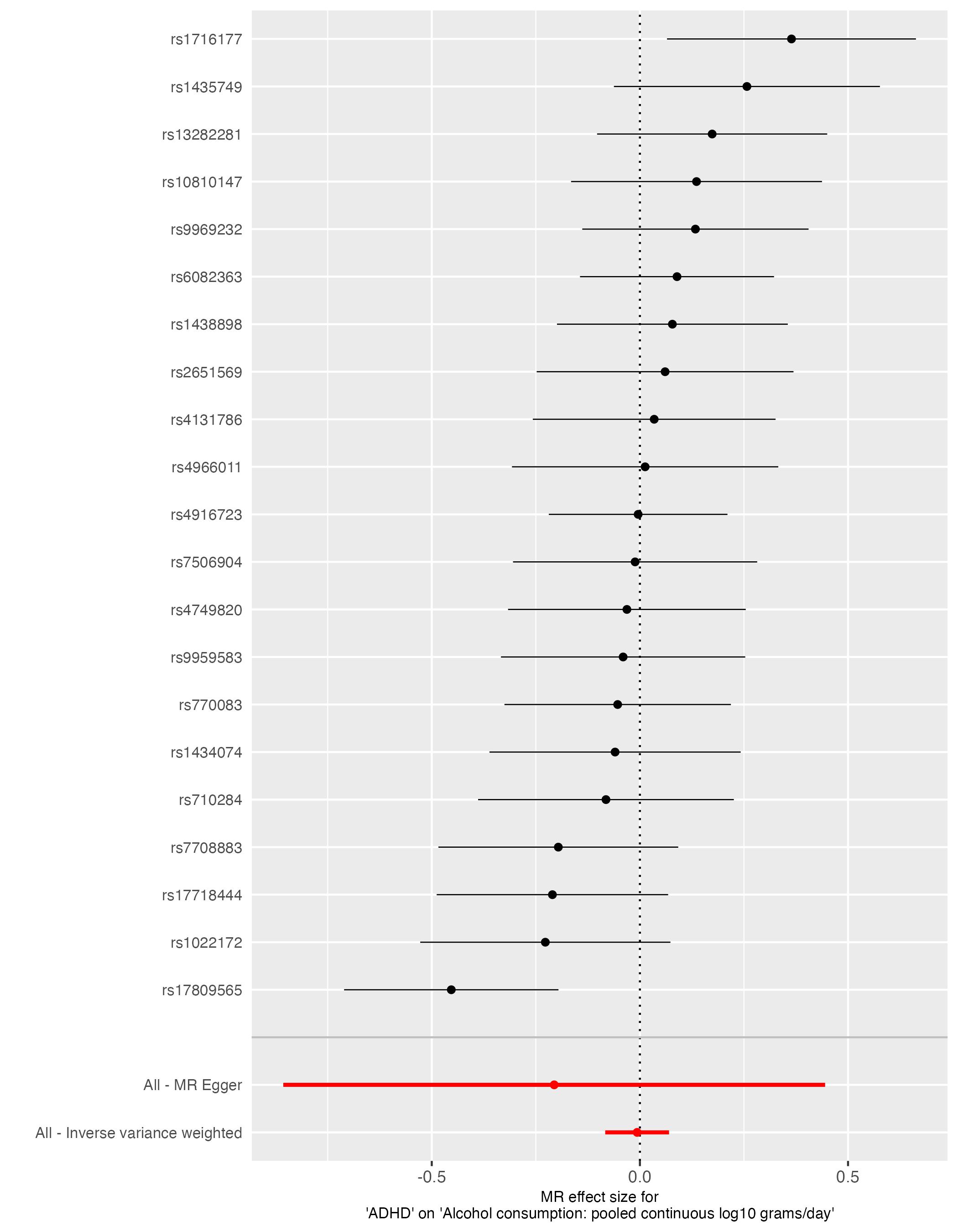

Supplementary Figure 8. Scatter plot analysis from UVMR of attention deficit hyperactivity disorder (ADHD) on alcohol use

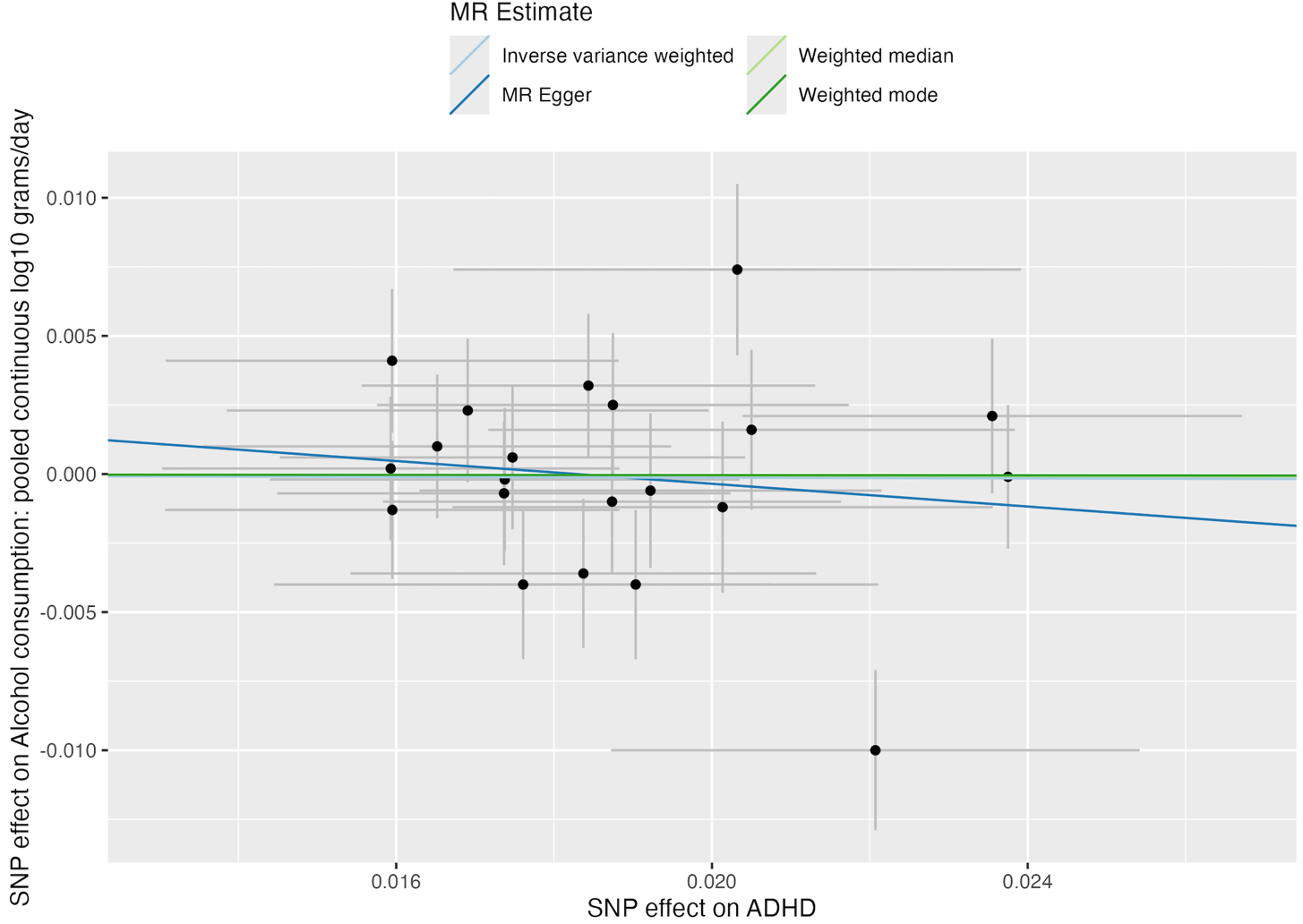

Supplementary Figure 9. Single SNP analysis from UVMR of depression on alcohol use

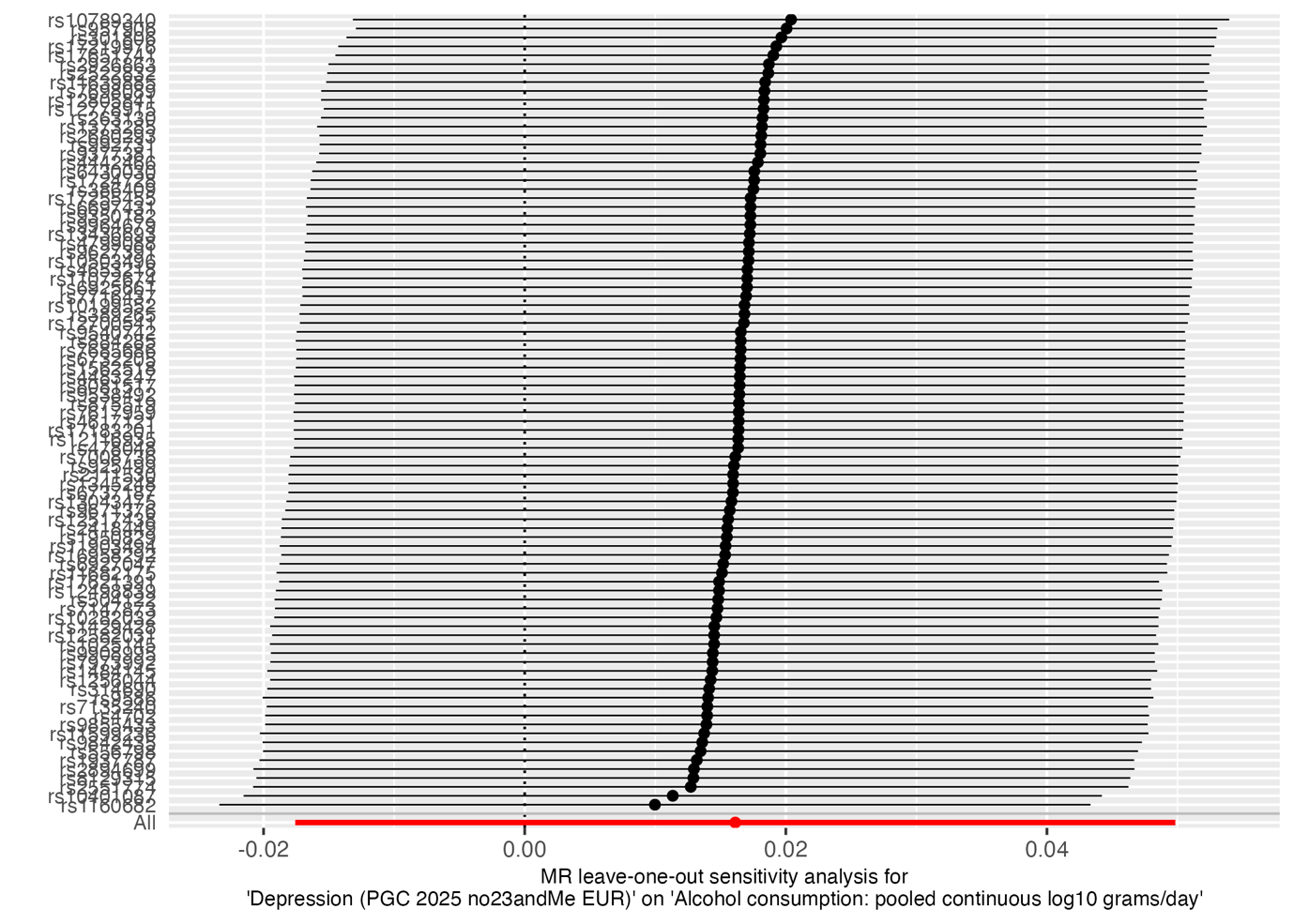

Supplementary Figure 10. Funnel plot analysis from UVMR of depression on alcohol use

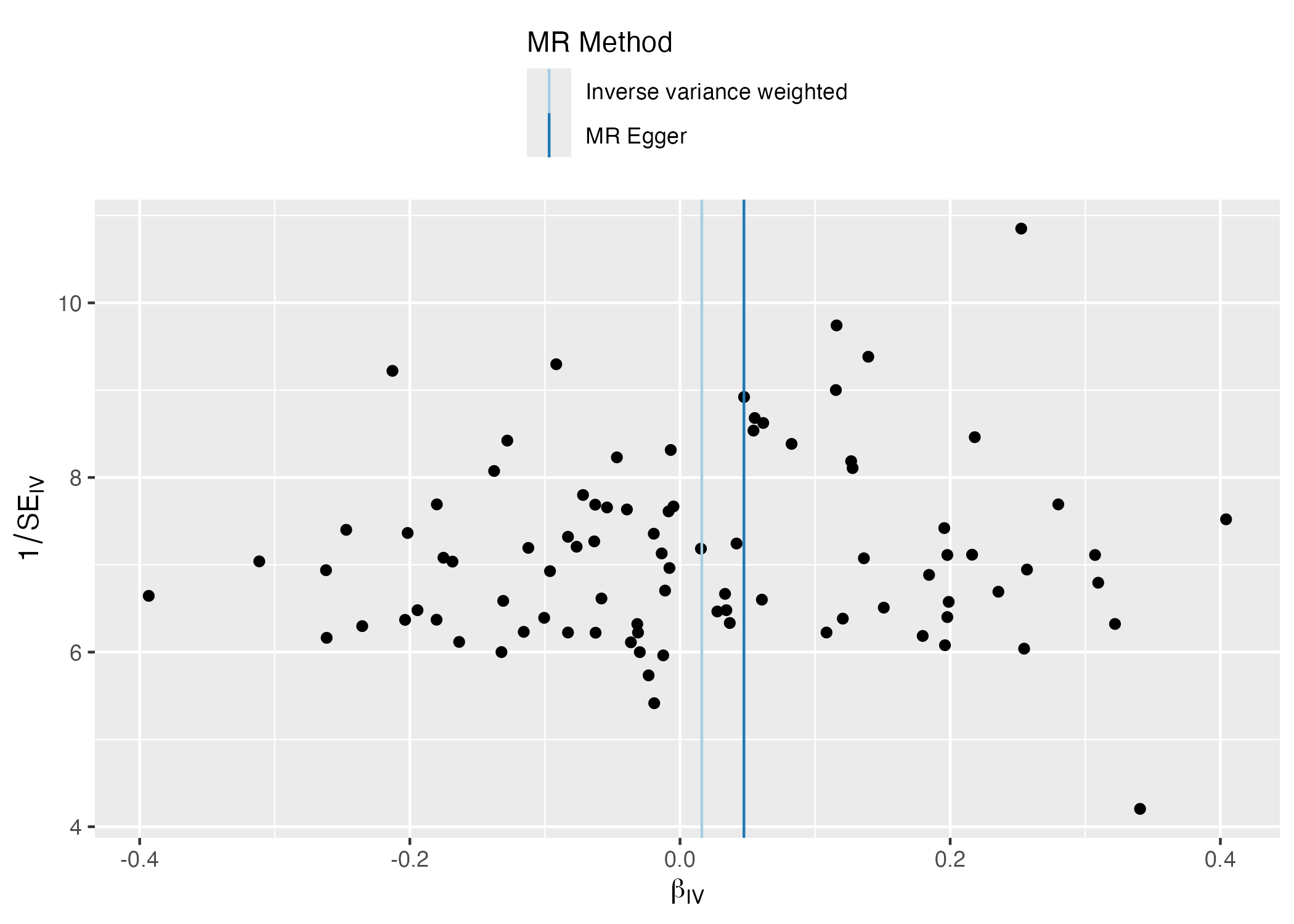

Supplementary Figure 11. Forest plot analysis from UVMR of depression on alcohol use

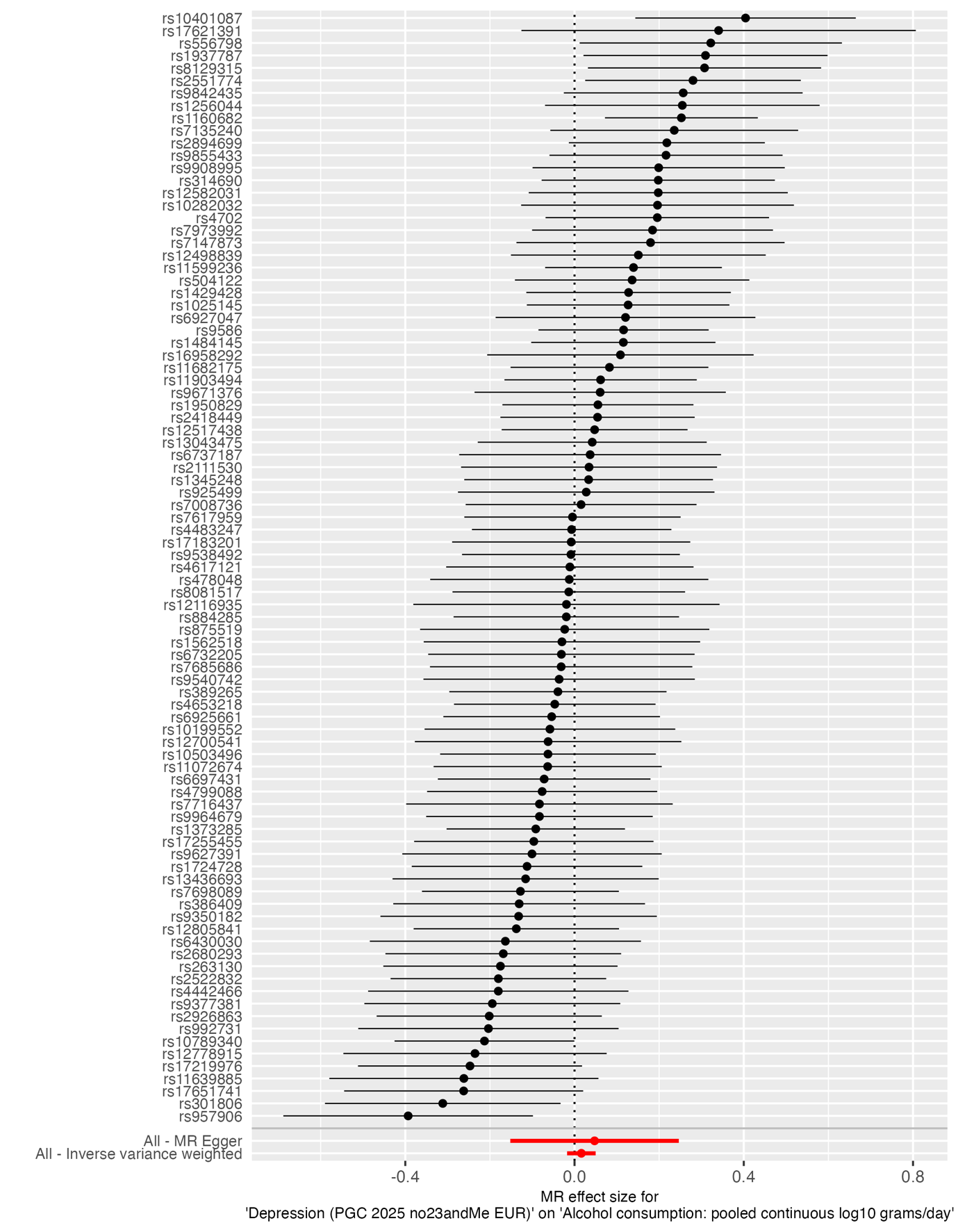

Supplementary Figure 12. Scatter plot analysis from UVMR of depression on alcohol use

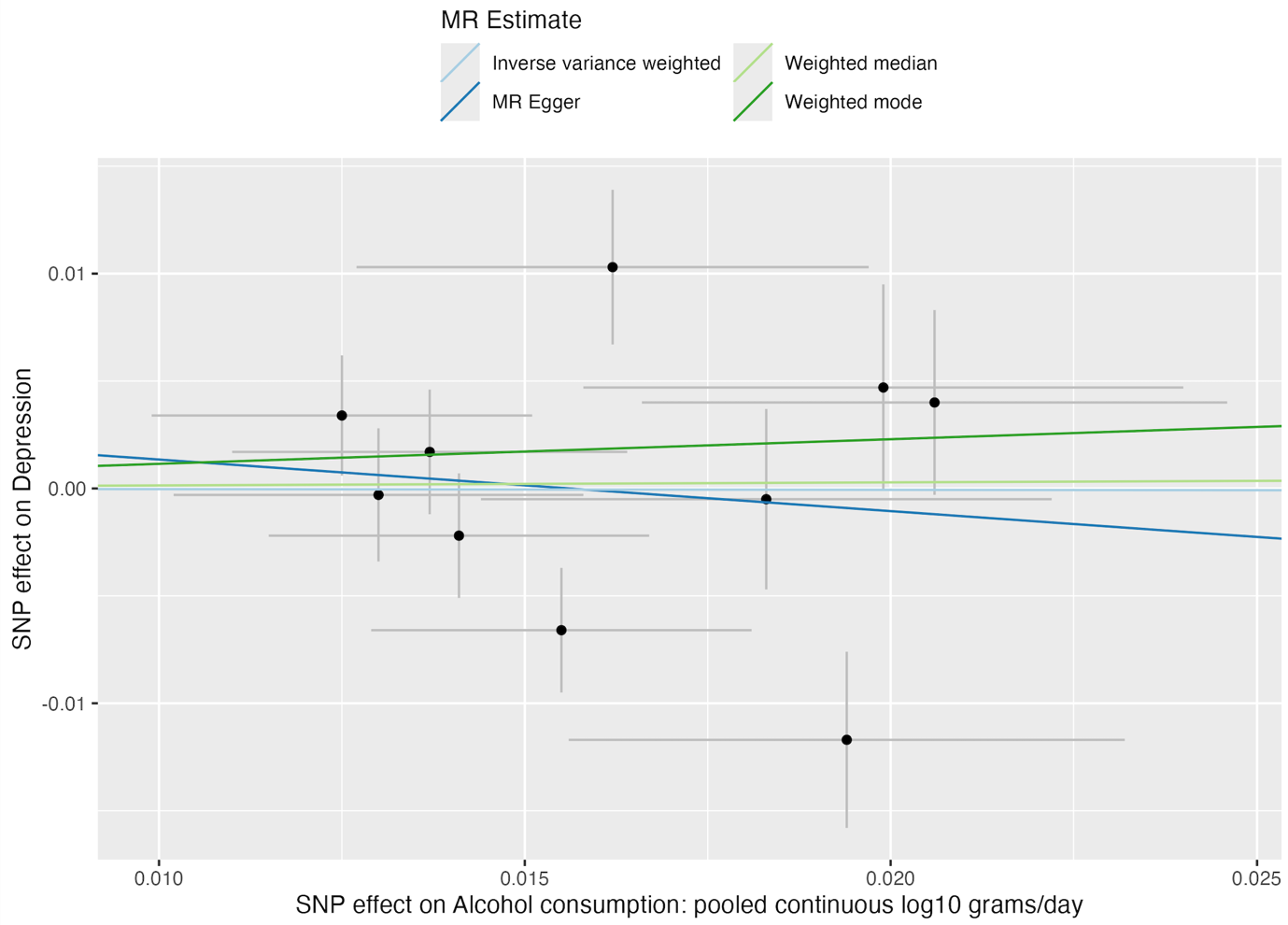

Supplementary Figure 13. Single SNP analysis from bidirectional UVMR of alcohol use on autism

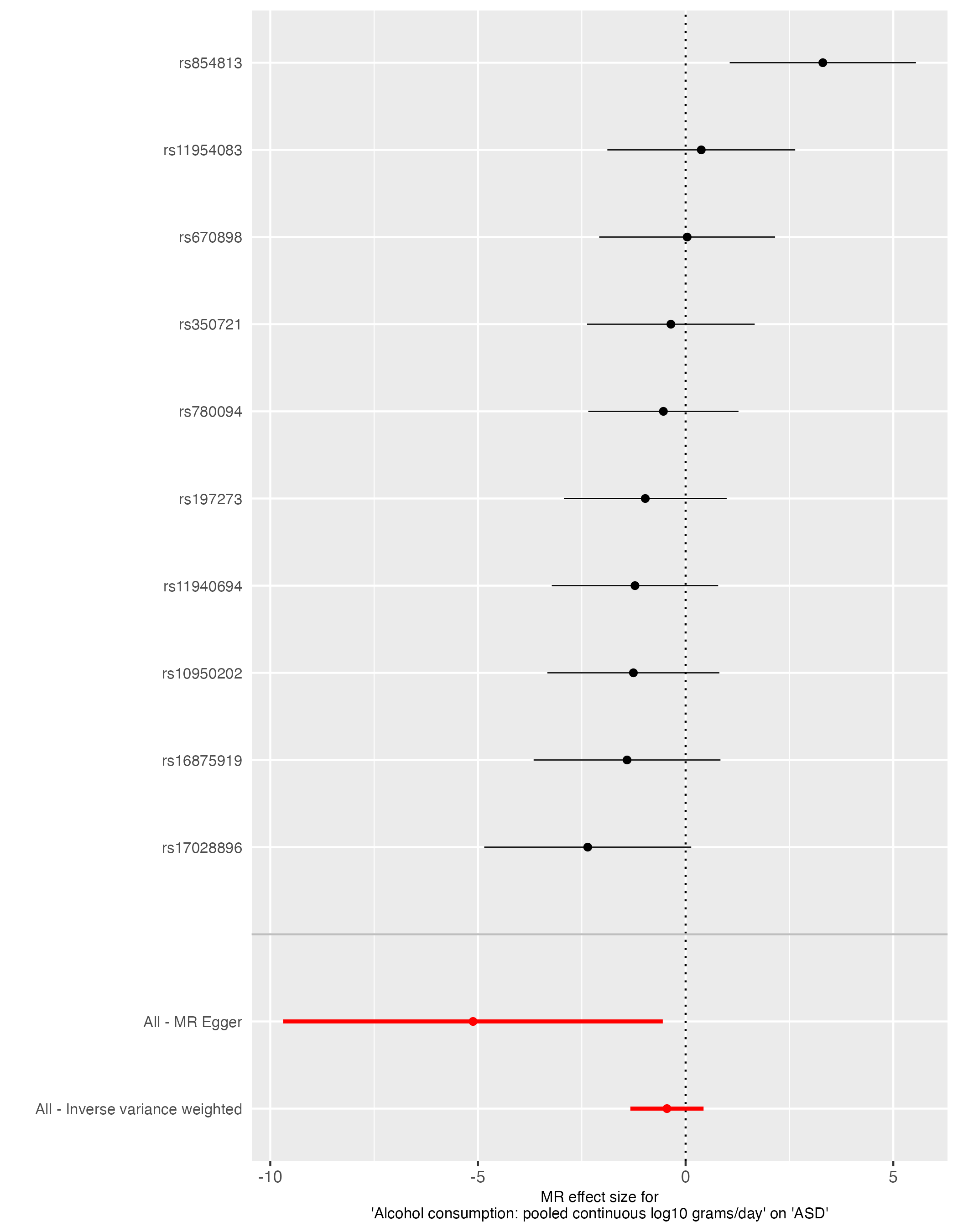

Supplementary Figure 14. Funnel plot from bidirectional UVMR of alcohol use on autism

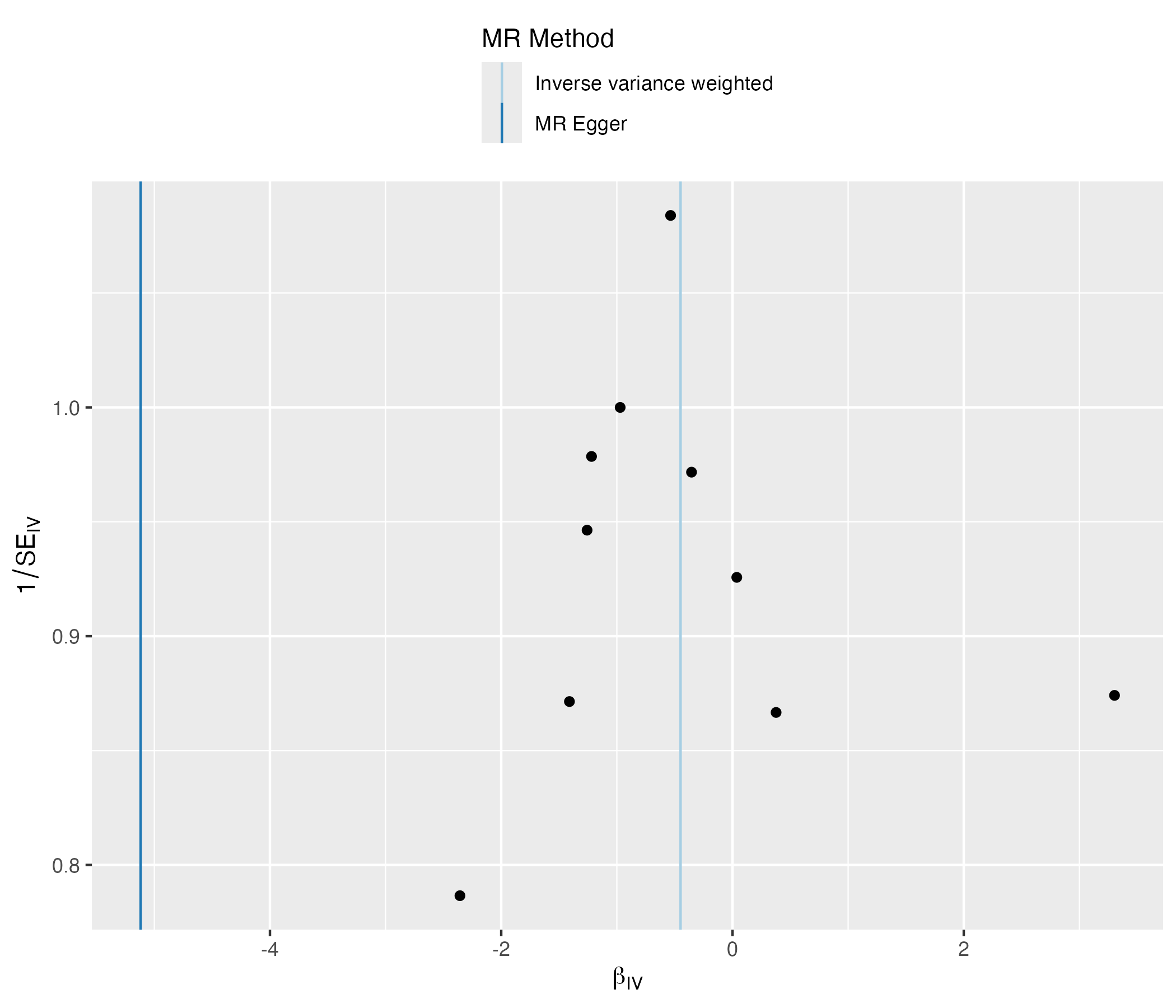

Supplementary Figure 15. Forest plot analysis from bidirectional UVMR of alcohol use on autism

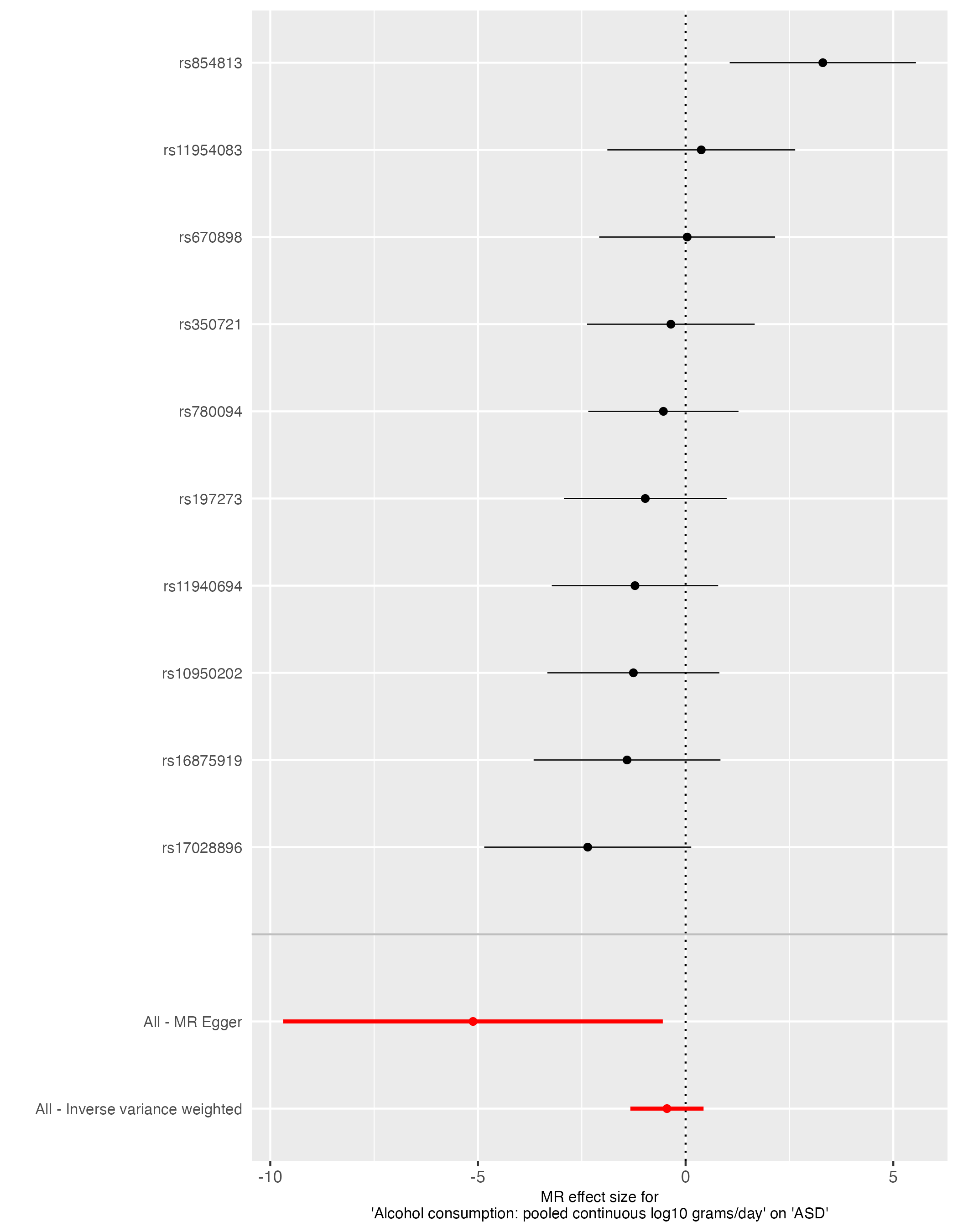

Supplementary Figure 16. Scatter plot analysis from bidirectional UVMR of alcohol use on autism

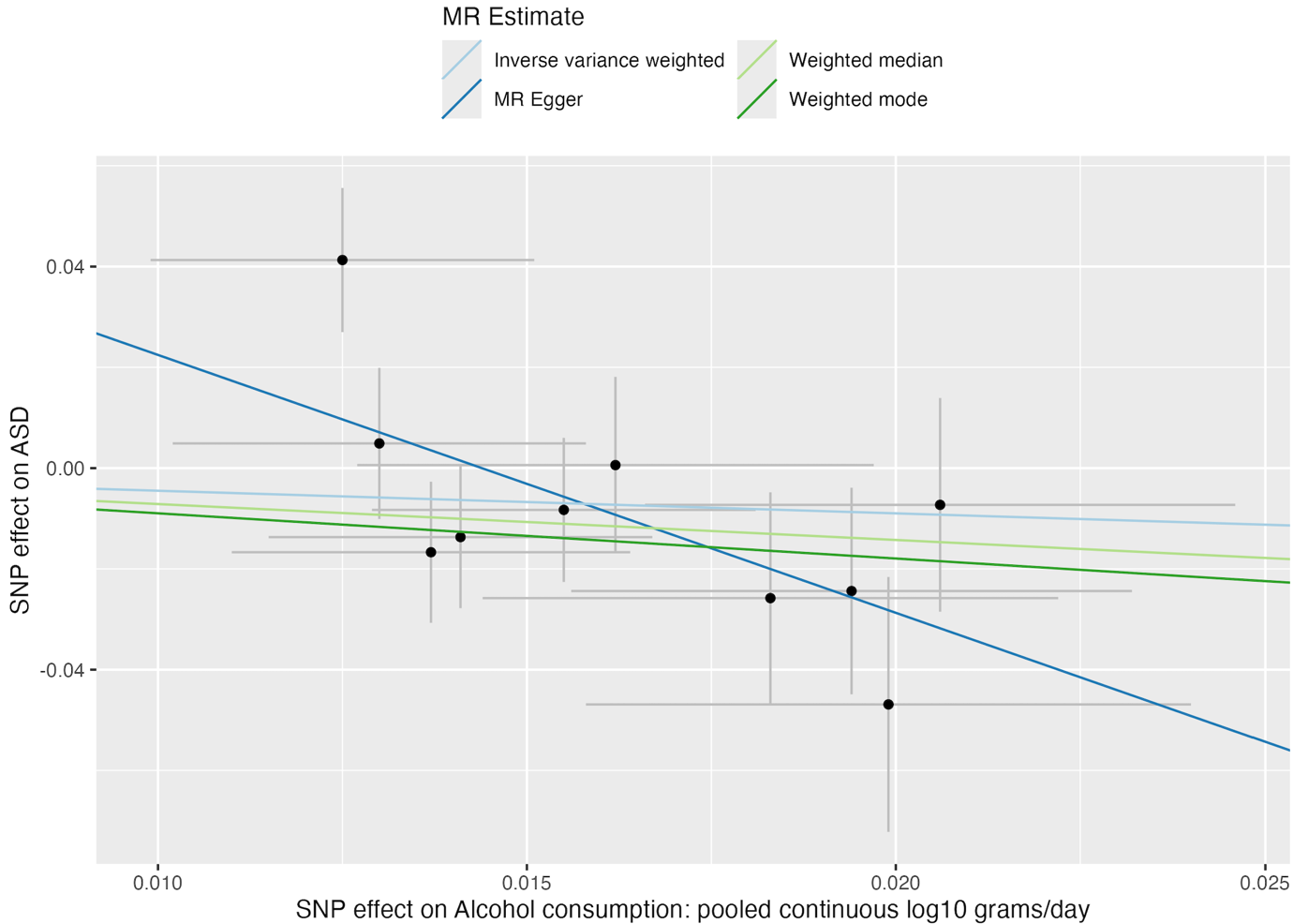

Supplementary Figure 17. Single SNP analysis from bidirectional UVMR of alcohol use on attention deficit hyperactivity disorder (ADHD)

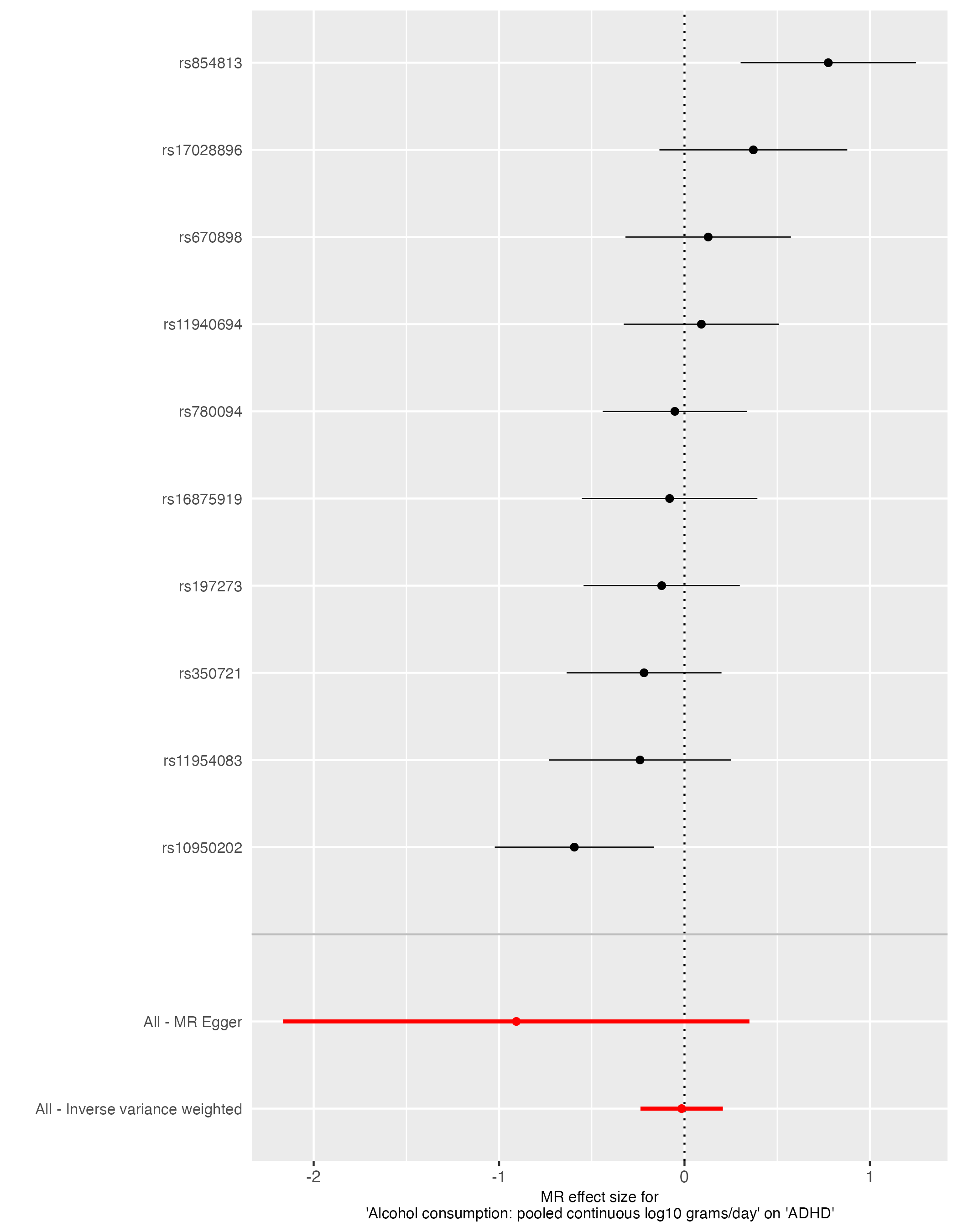

Supplementary Figure 18. Funnel plot analysis from bidirectional UVMR of alcohol use on attention deficit hyperactivity disorder (ADHD)

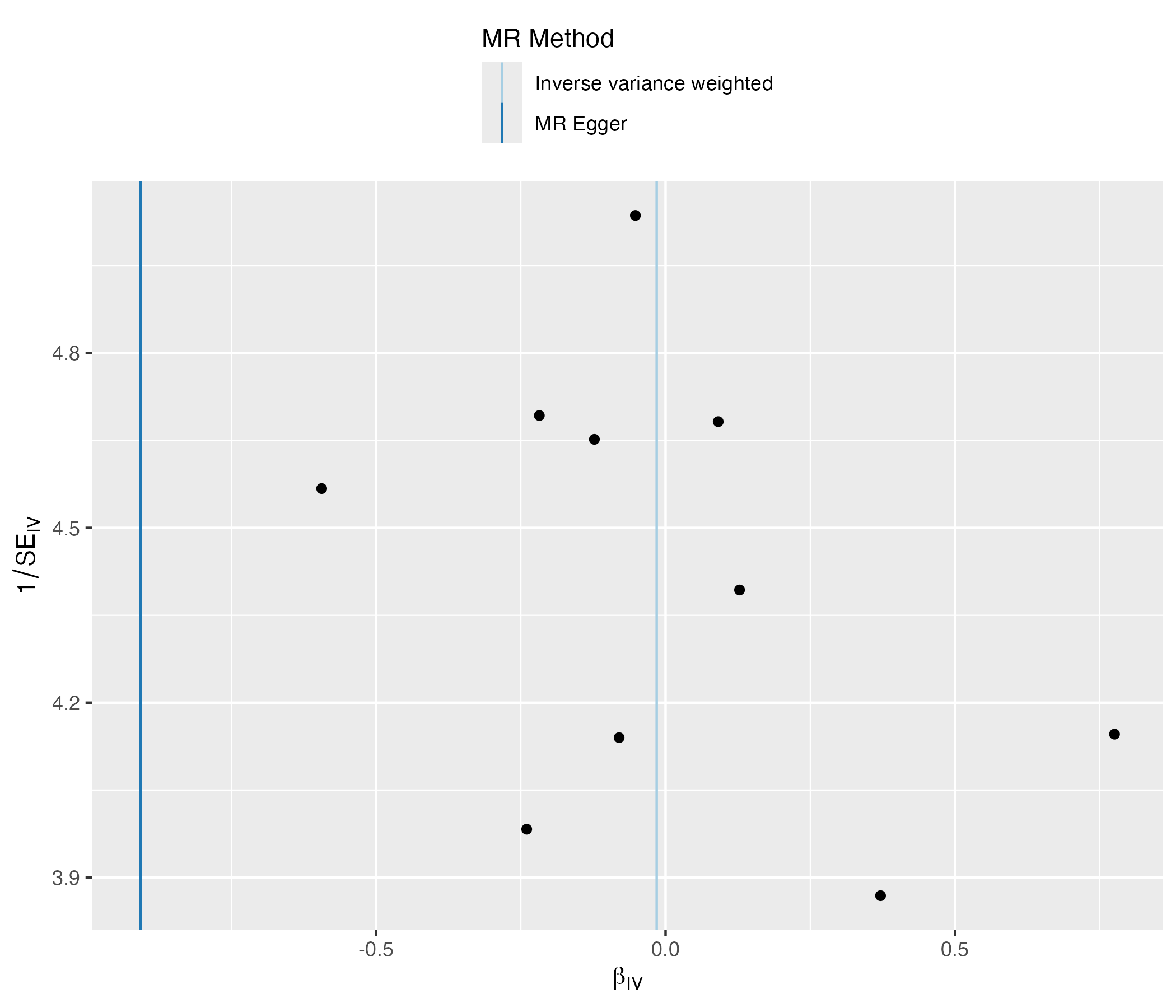

Supplementary Figure 19. Forest plot analysis from bidirectional UVMR of alcohol use on attention deficit hyperactivity disorder (ADHD)

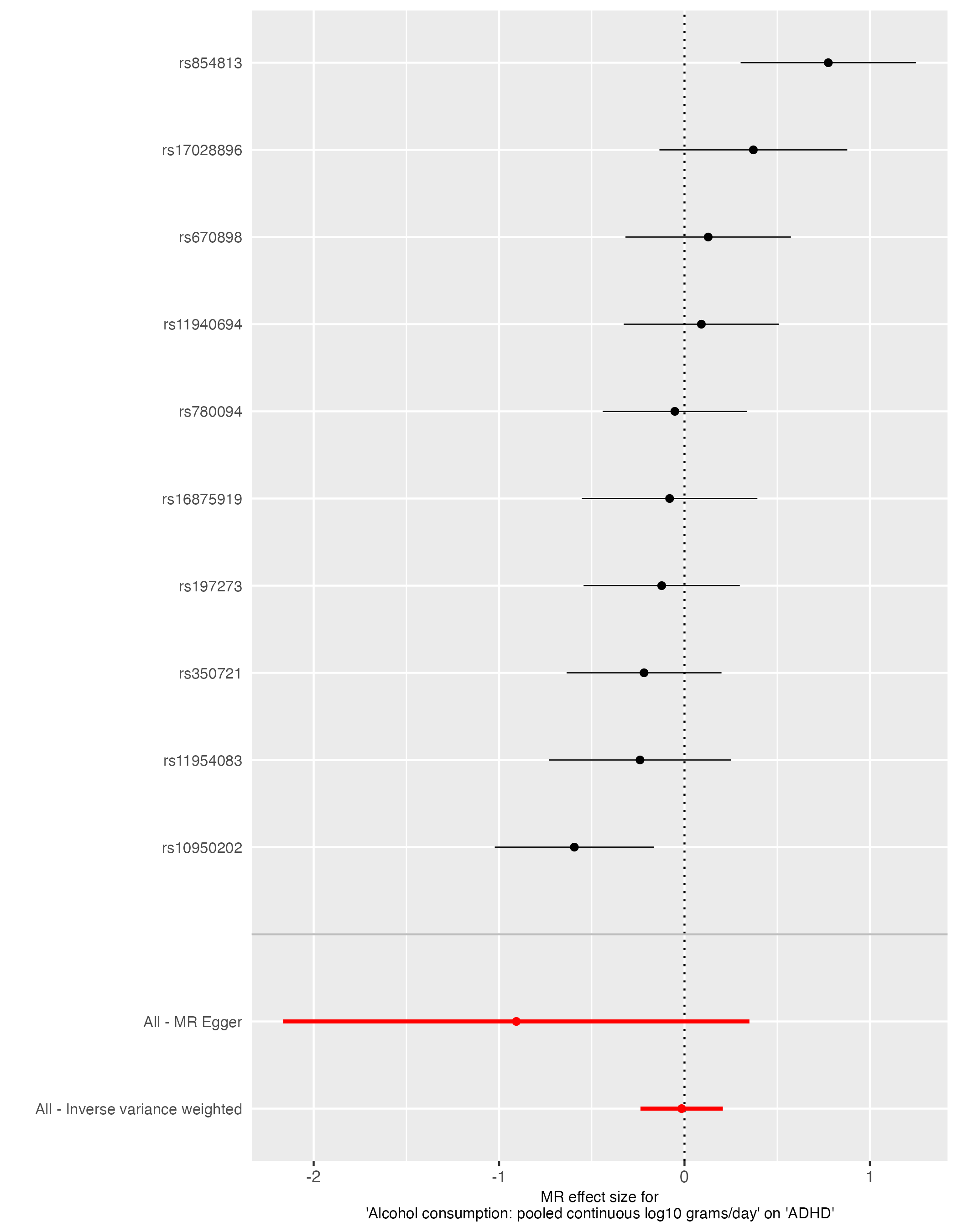

Supplementary Figure 20. Scatter plot analysis from bidirectional UVMR of alcohol use on attention deficit hyperactivity disorder (ADHD)

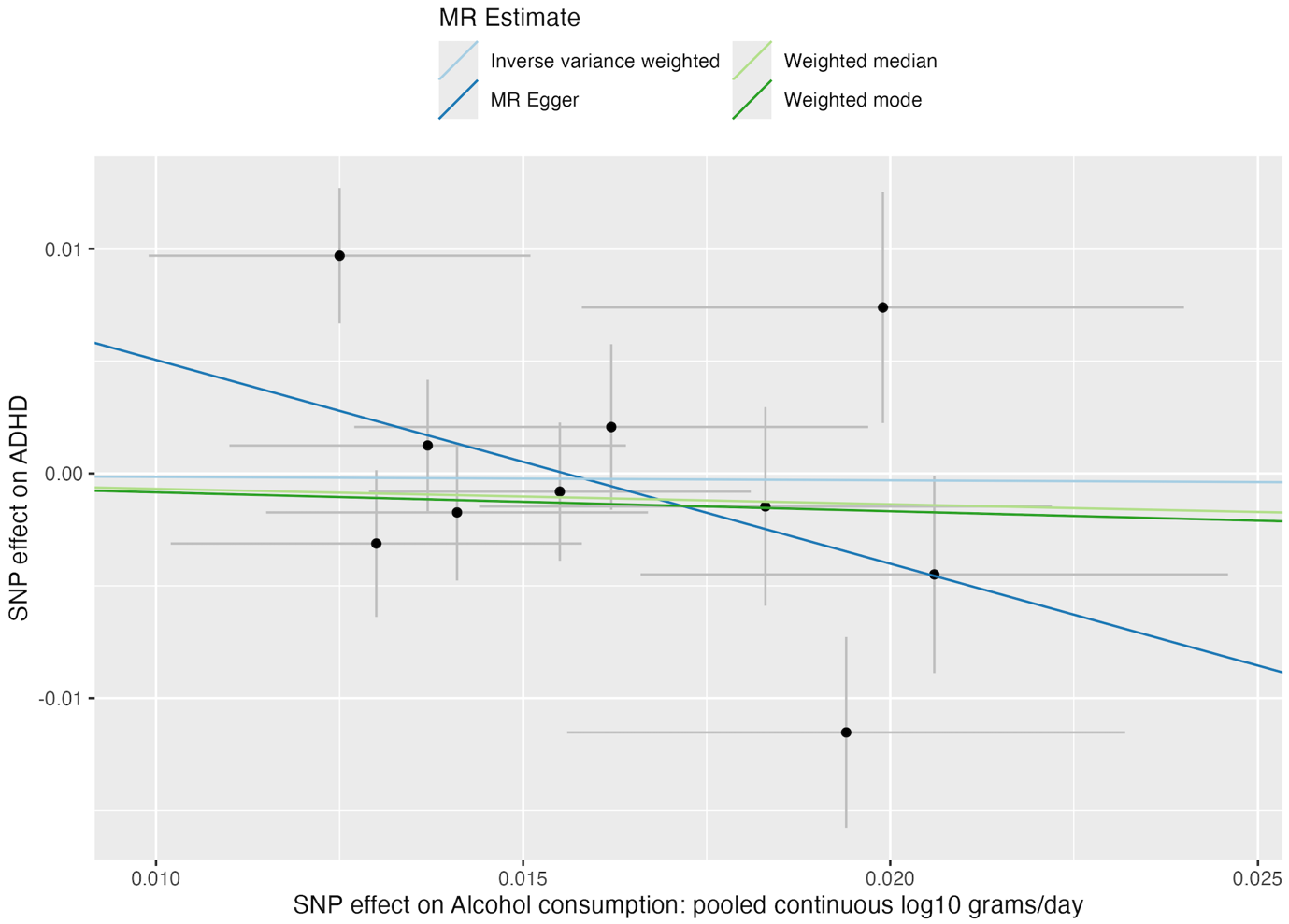

Supplementary Figure 21. Single SNP analysis from bidirectional UVMR of alcohol use on depression

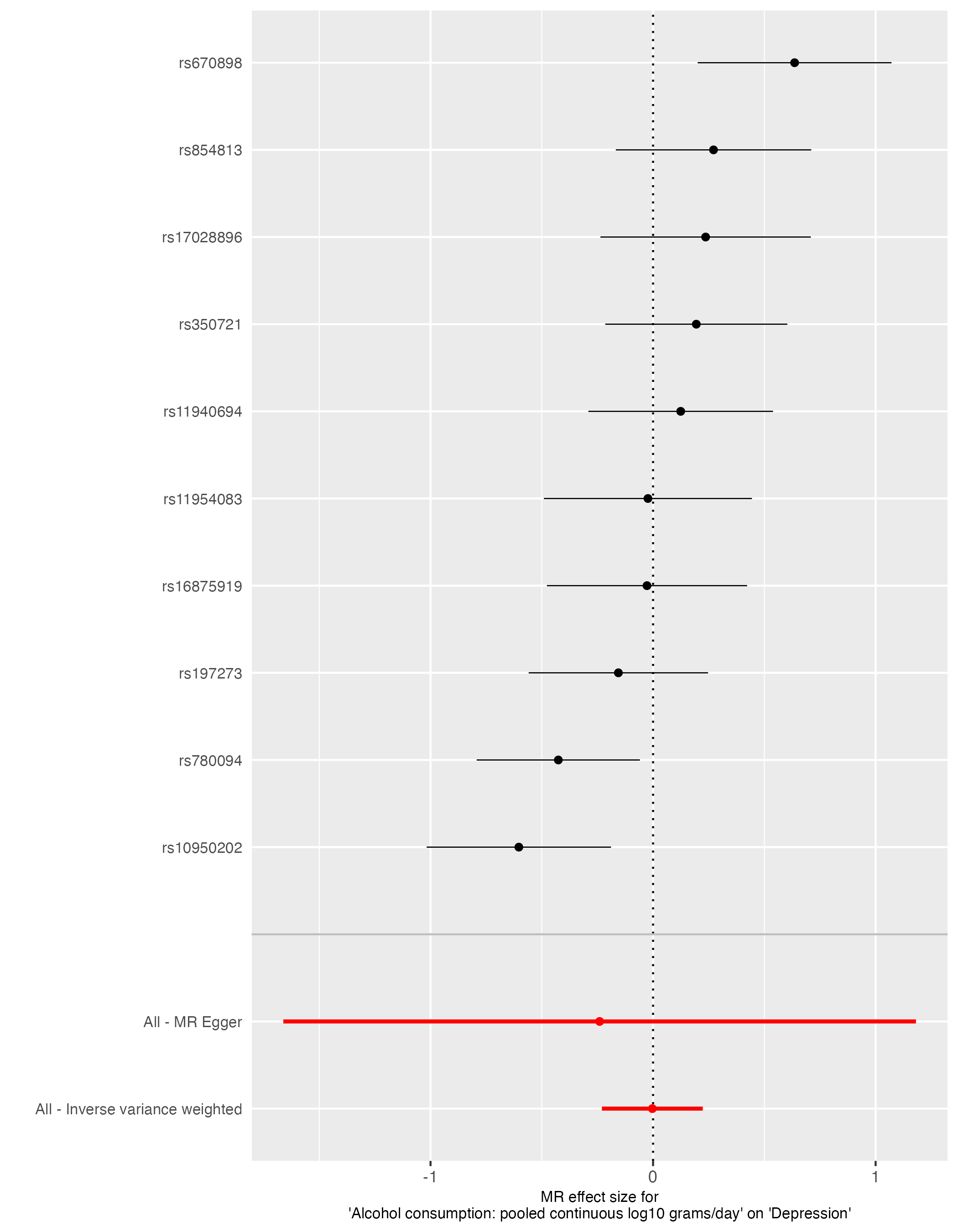

Supplementary Figure 22. Funnel plot analysis from bidirectional UVMR of alcohol use on depression

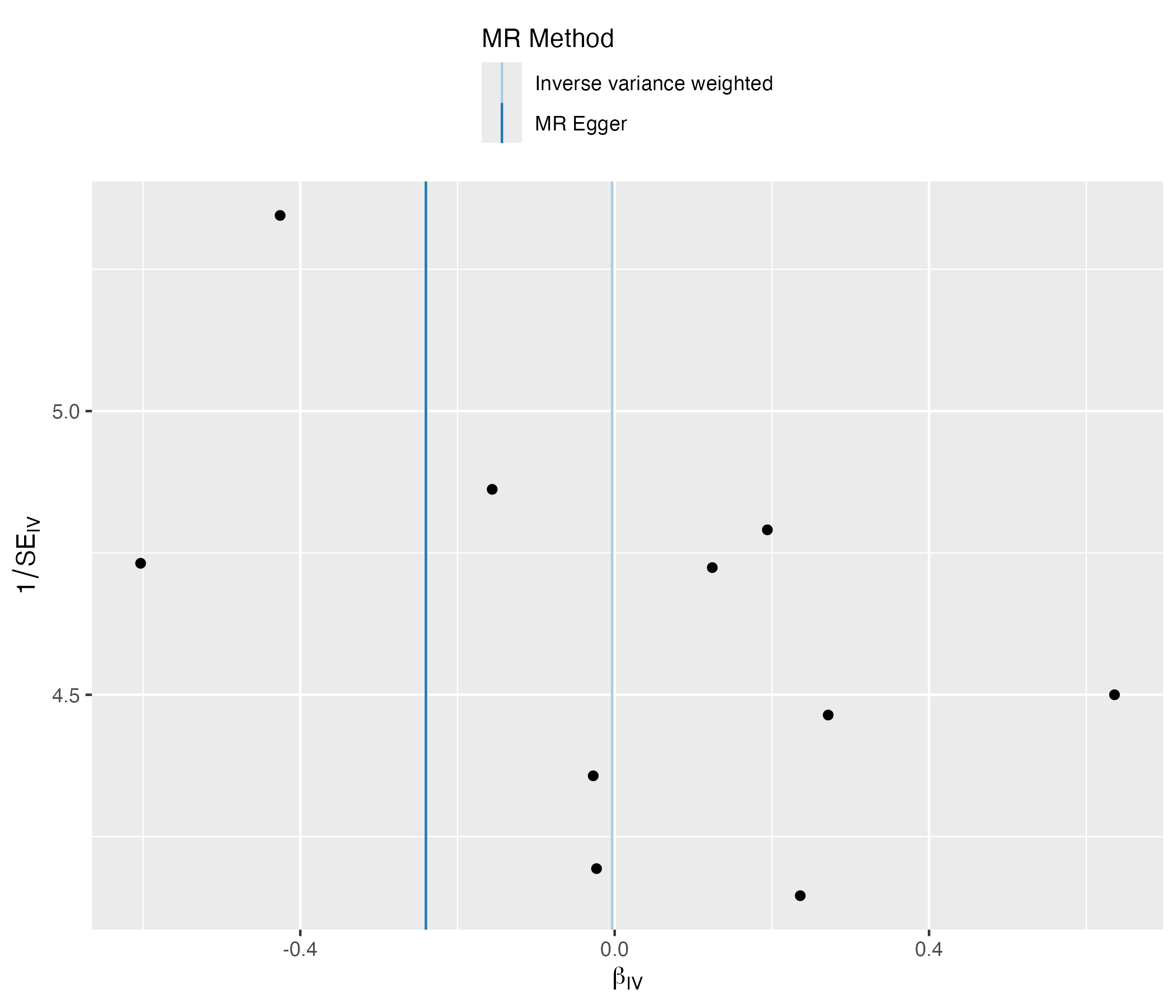

Supplementary Figure 23. Forest plot analysis from bidirectional UVMR of alcohol use on depression

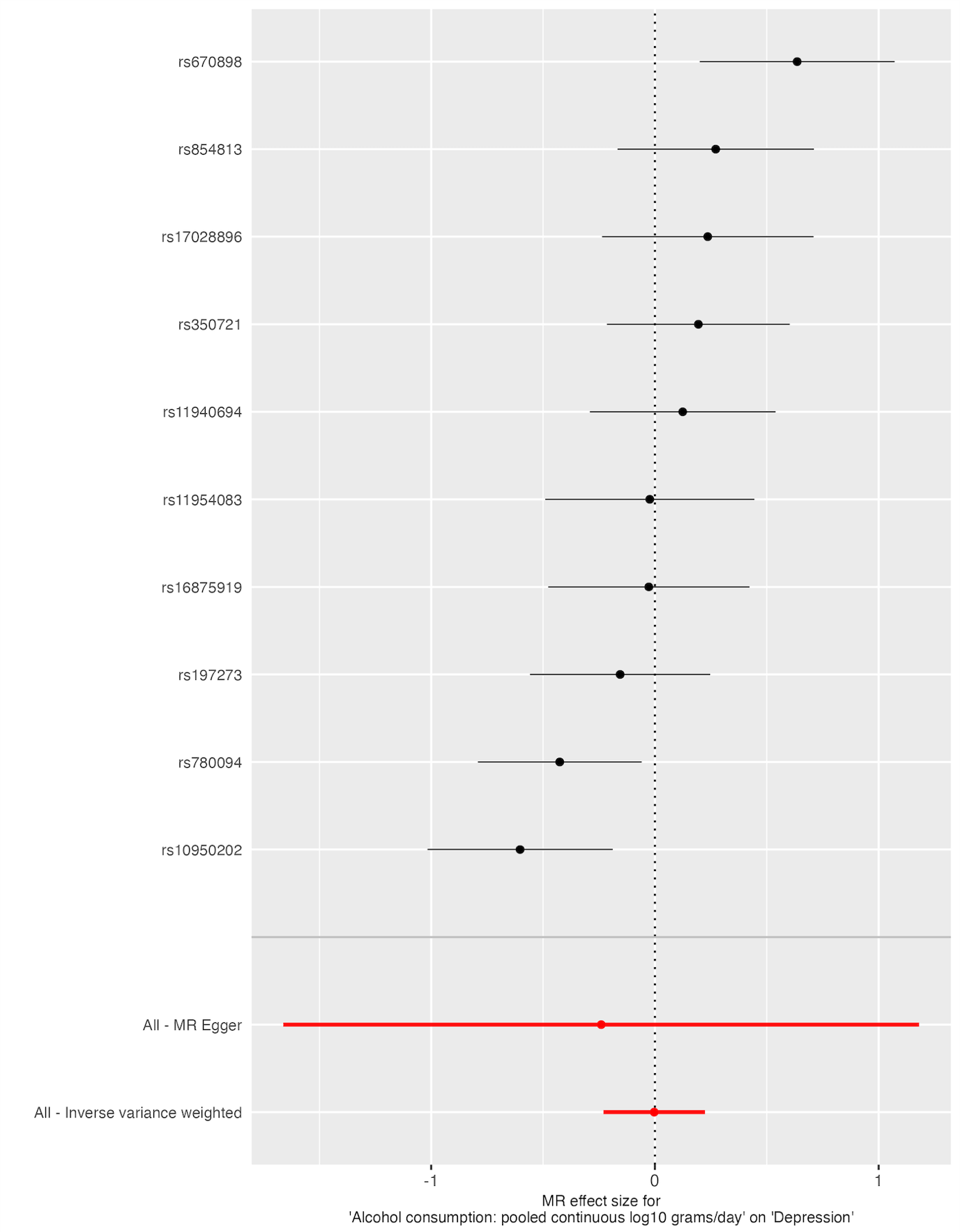

Supplementary Figure 24. Scatter plot analysis from bidirectional UVMR of alcohol use

on depression

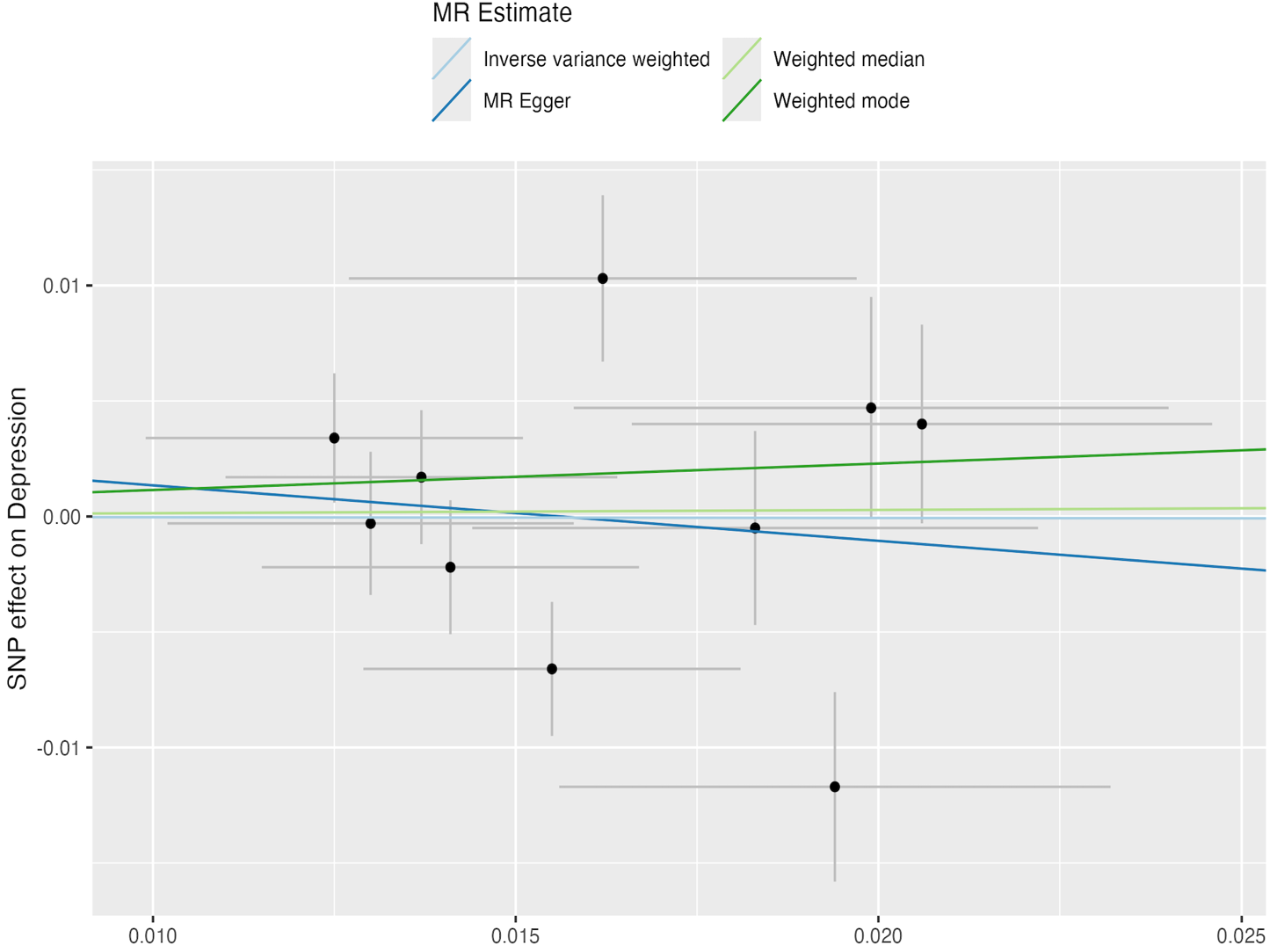

**The following figures represent analysis results after Steiger filtering. Please note, analyses including instruments for autism and ADHD remained unchanged, so figures are given for depression only.**

Supplementary Figure 25. Single SNP analysis from UVMR of depression on alcohol use

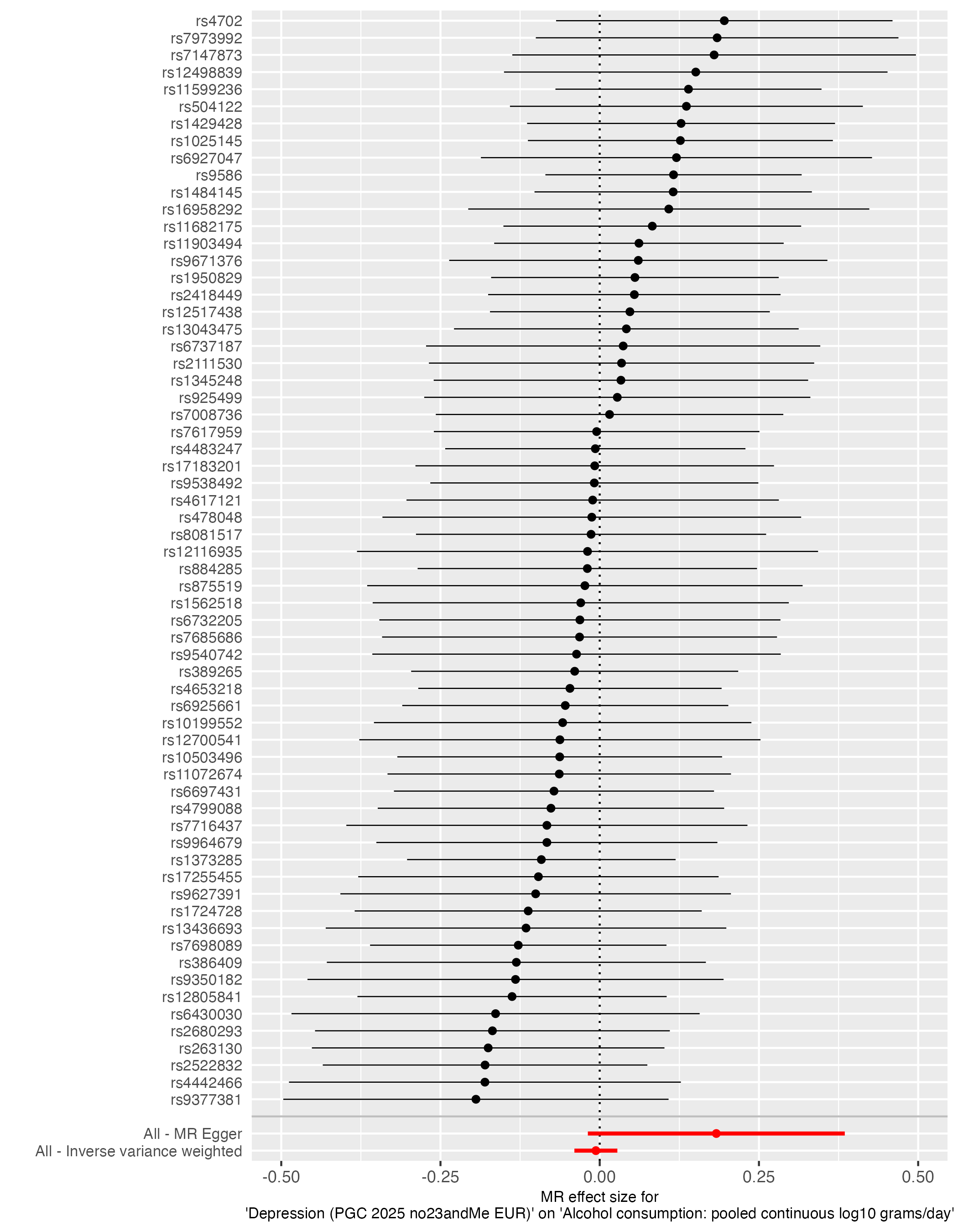

Supplementary Figure 26. Funnel plot analysis from UVMR of depression on alcohol use

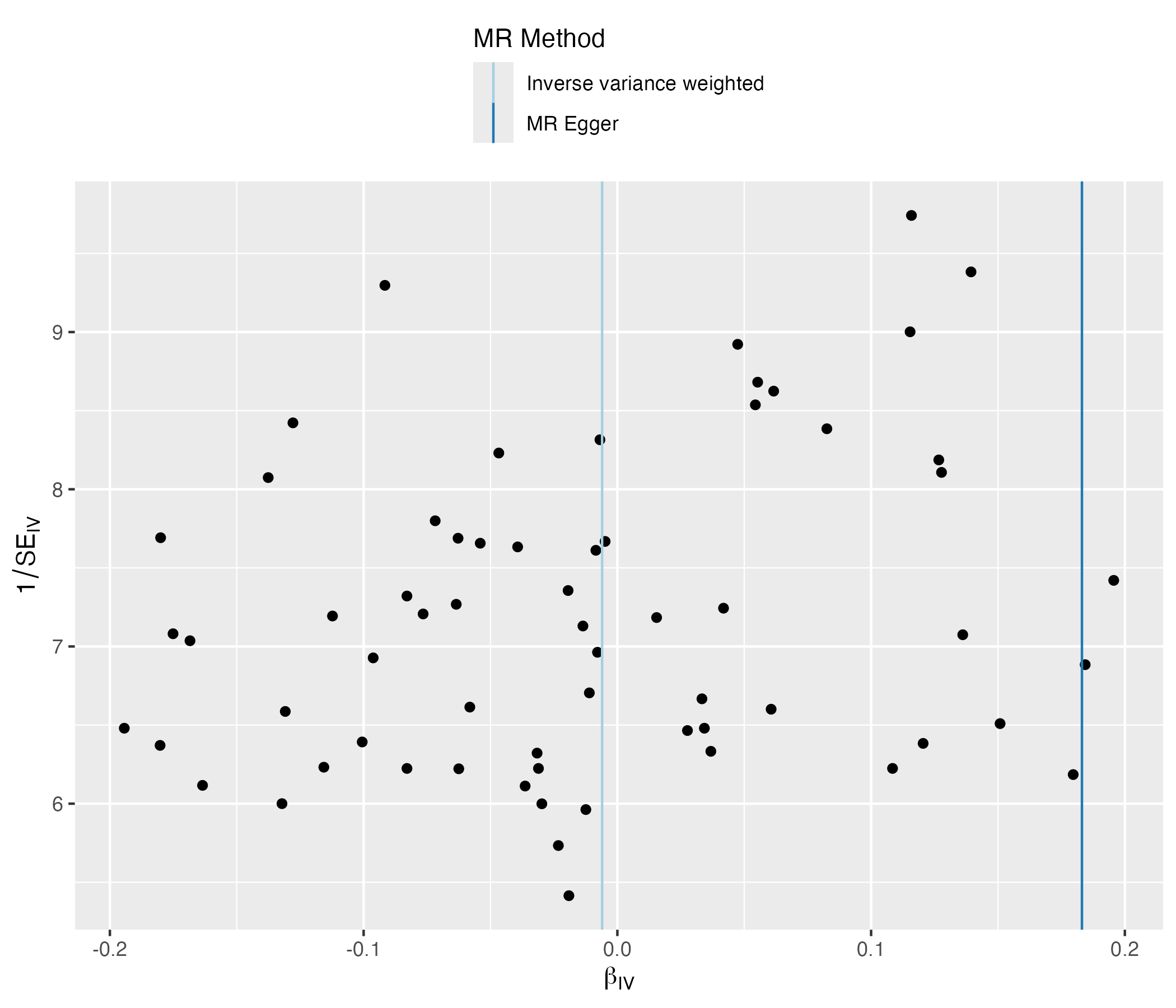

Supplementary Figure 27. Forest plot analysis from UVMR of depression on alcohol use

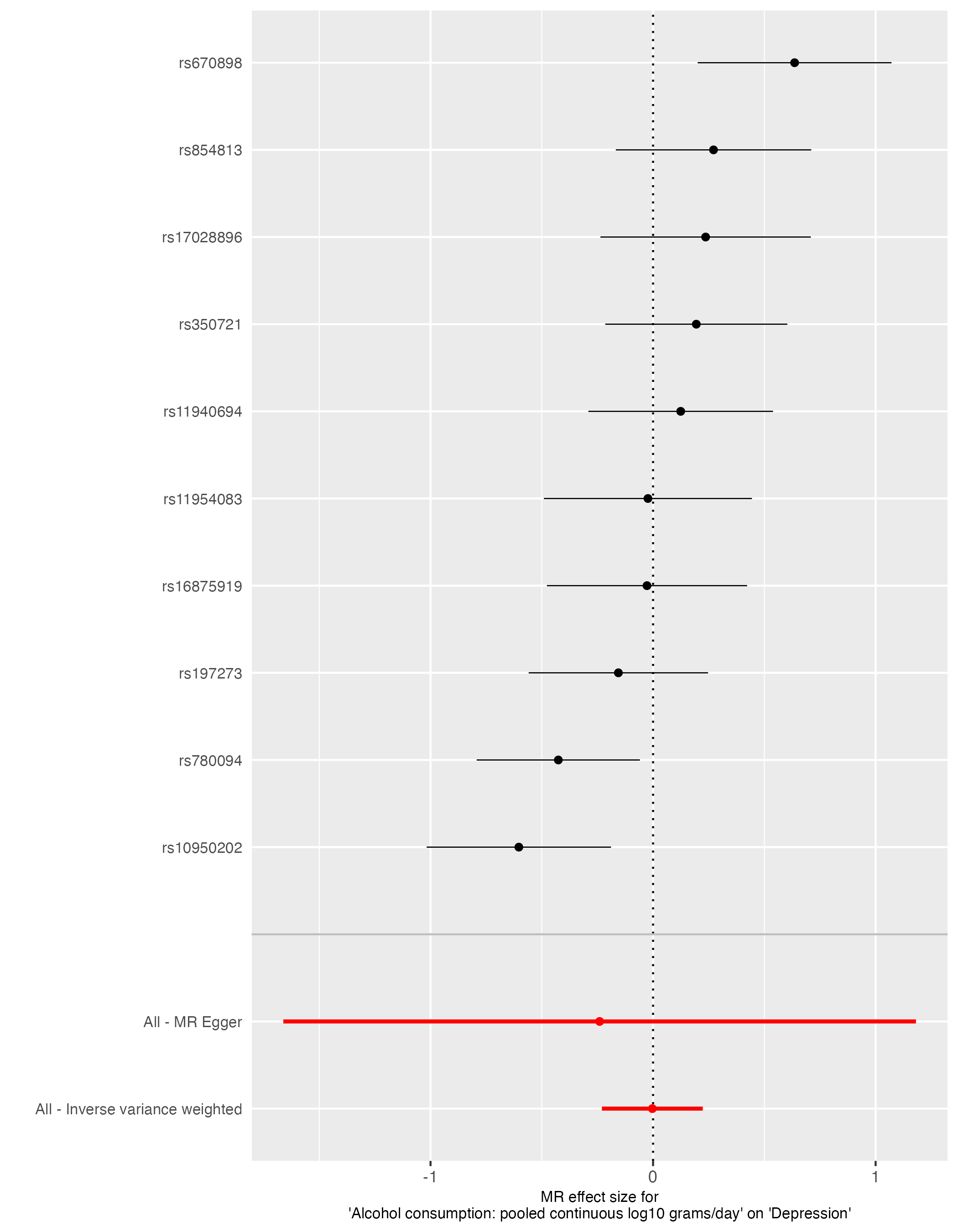

Supplementary Figure 28. Scatter plot analysis from UVMR of depression on alcohol use

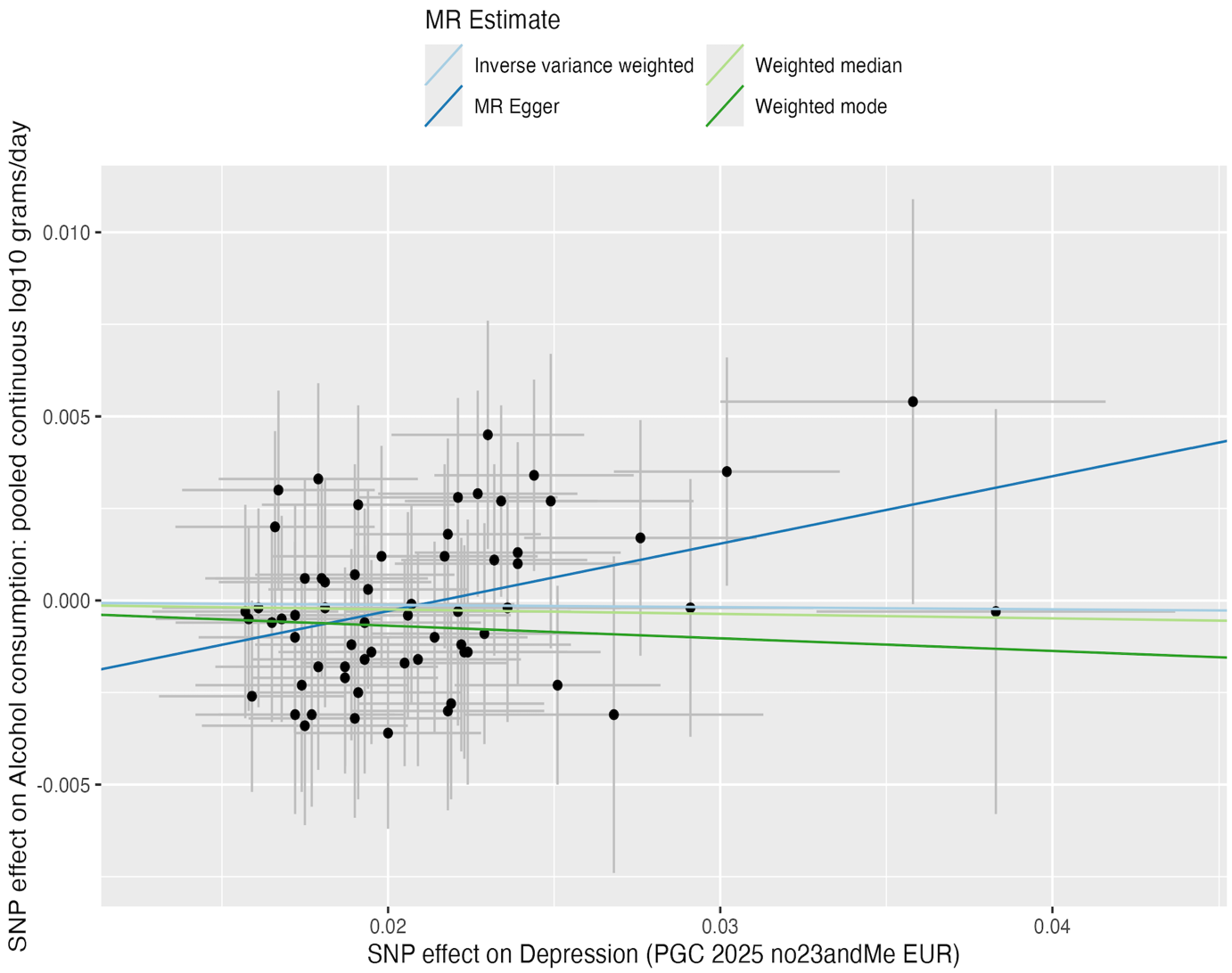

Supplementary Figure 29. Single SNP analysis from bidirectional UVMR of alcohol use on depression

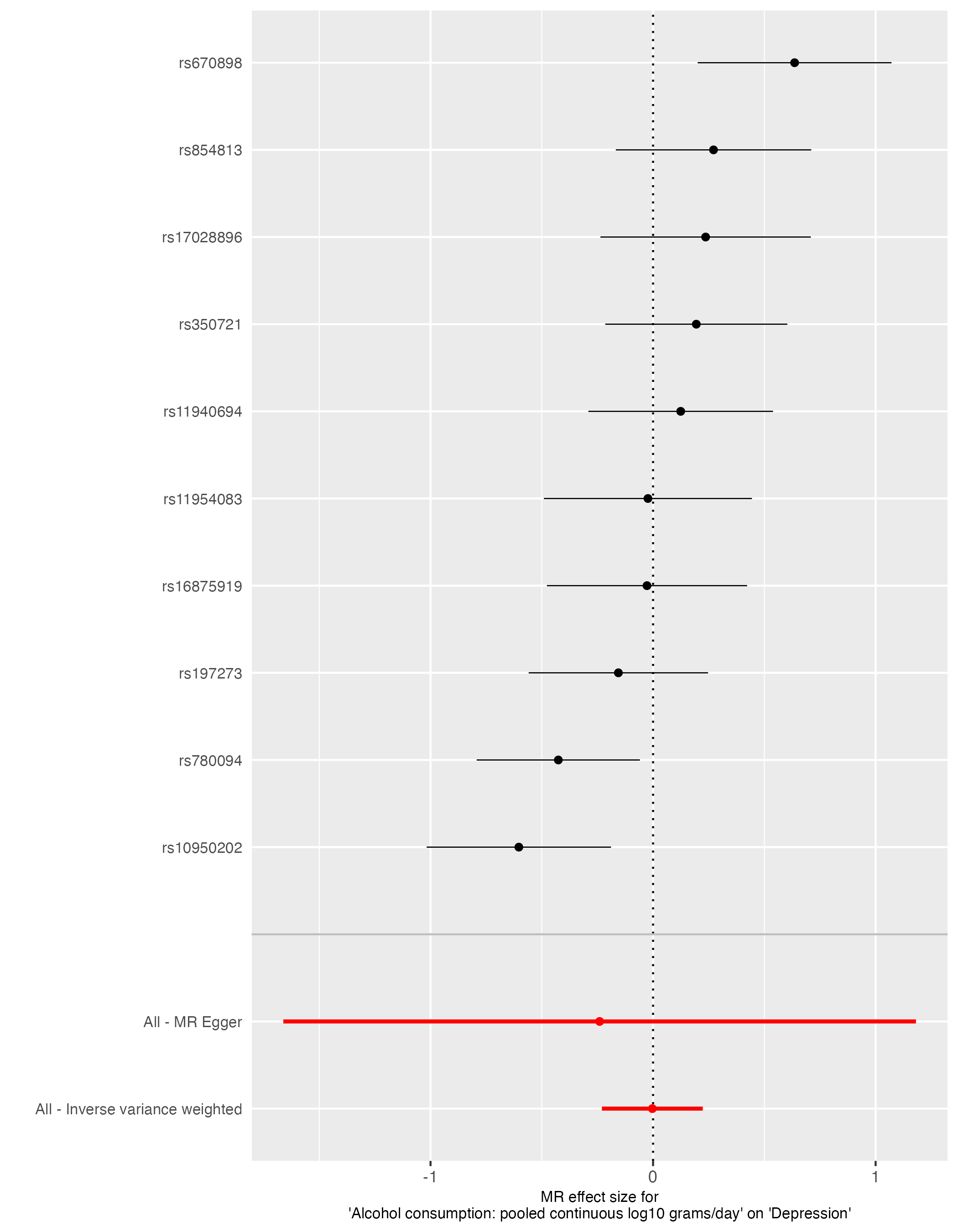

Supplementary Figure 30. Funnel plot analysis from bidirectional UVMR of alcohol use on depression

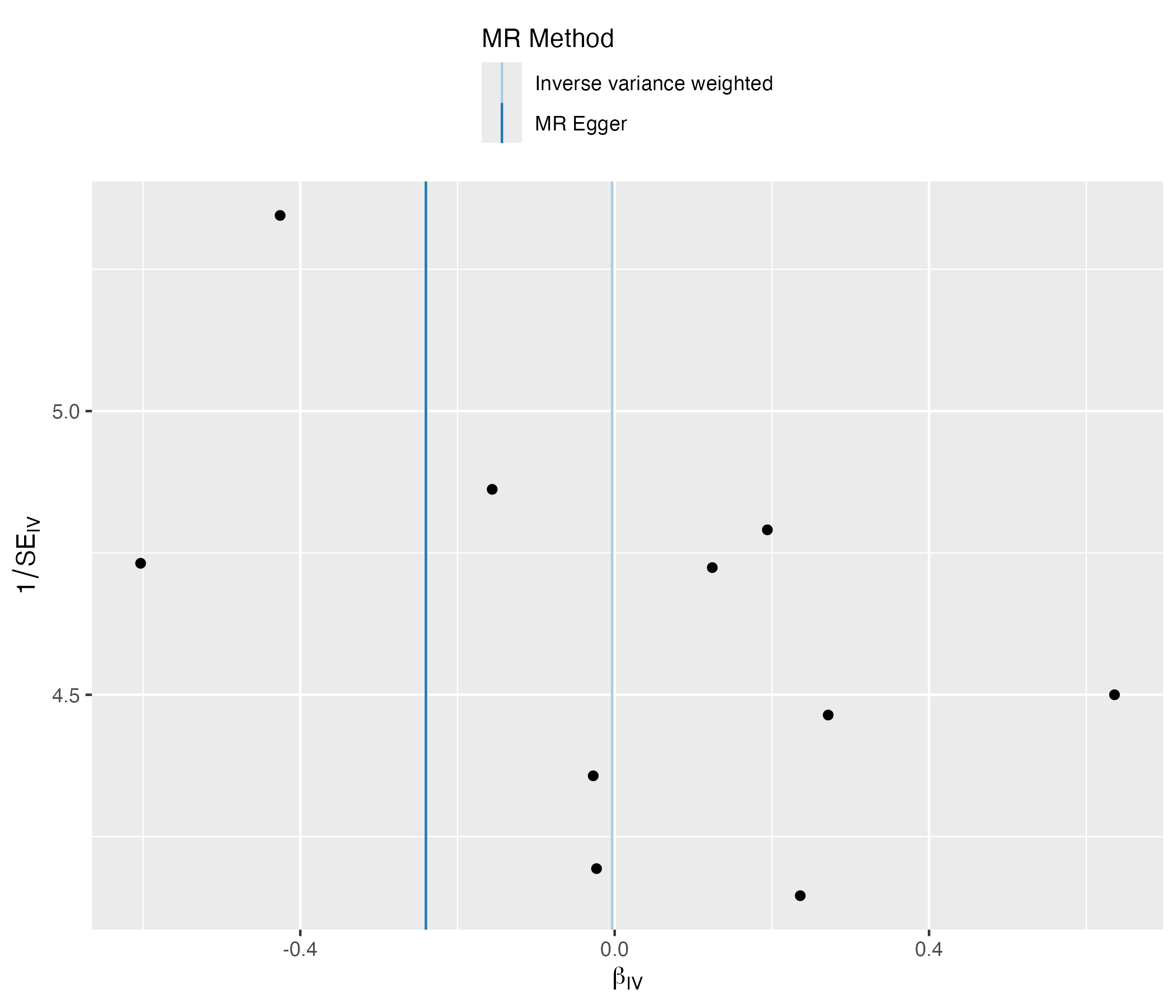

Supplementary Figure 31. Forest plot analysis from bidirectional UVMR of alcohol use on depression

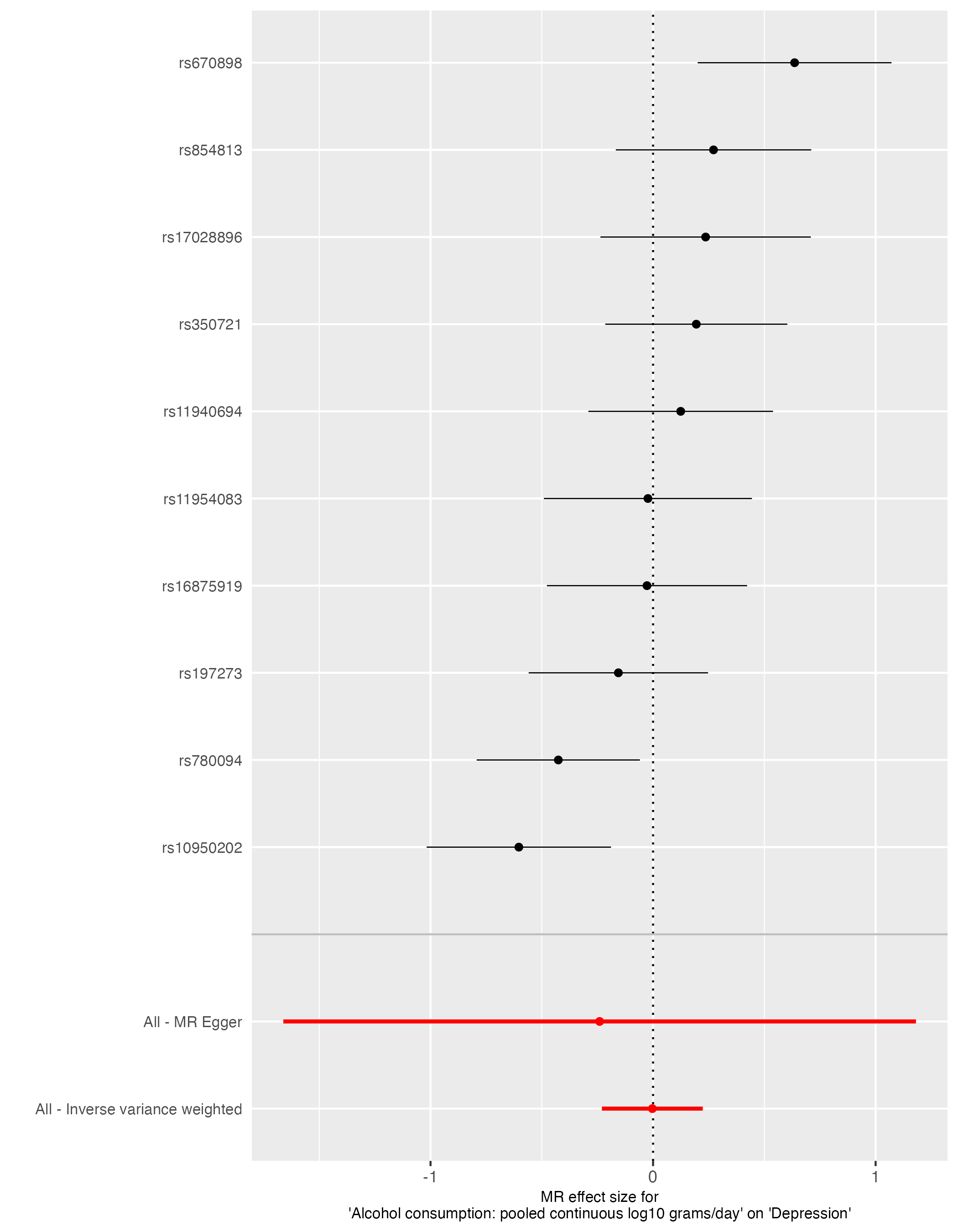

Supplementary Figure 32. Scatter plot analysis from bidirectional UVMR of alcohol use on depression

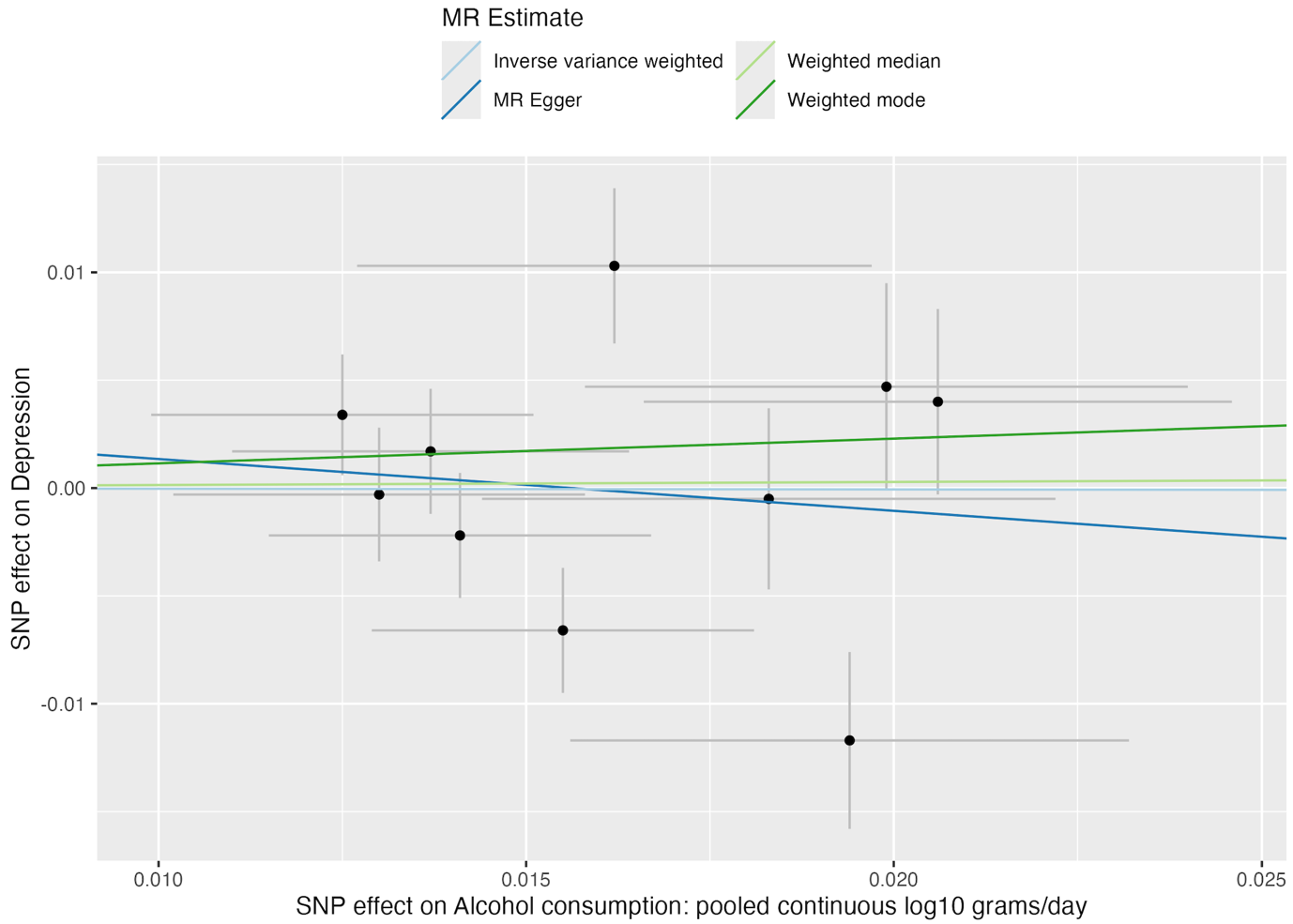
