## Supplementary Tables for "Does genetic liability for autism influence alcohol use?"

**PGS analyses**

*Supplementary Table S1*. List of p value thresholds used in calculation of polygenic score for autism

| **P value** | **Threshold** |
| --- | --- |
| p5e-8 | 0.00000005 |
| p1e-6 | 0.000001 |
| p1e-4 | 0.001 |
| p0.01 | 0.01 |
| p0.05 | 0.05 |
| p0.1 | 0.1 |
| p0.5 | 0.5 |
| p1 | 1 |

*Supplementary Table S2*. Response codes for phenotypic behaviours measured in UK Biobank

| **Variable** | **N** | **Question** | **Code** |
| --- | --- | --- | --- |
| Average monthly red wine intake* | 62,902 | "In an average MONTH, how many glasses of RED wine would you drink? (There are six glasses in an average bottle)" | 4407 |
| Average monthly champagne plus white intake* | 62,902 | "In an average MONTH, how many glasses of WHITE wine would you drink? (There are six glasses in an average bottle)" | 4418 |
| Average monthly beer plus cider intake* | 62,902 | "In an average MONTH, how many pints of beer or cider would you drink? (Include bitter, lager, stout, ale, Guinness)" | 4429 |
| Average monthly spirits intake* | 62,902 | "In an average MONTH, how many measures of spirits or liqueurs would you drink? (there are 25 standard measures in a normal sized bottle; spirits include drinks such as whisky, gin, rum, vodka, brandy)" | 4440 |
| Average monthly fortified wine intake* | 62,902 | "In an average MONTH, how many glasses of fortified wine would you drink? (There are 12 glasses in an average bottle) (Fortified wines include drinks such as sherry, port, vermouth)" | 4451 |
| Average monthly intake of other alcoholic drinks* | 62,902 | "In an average MONTH, how many glasses of other alcoholic drinks (such as alcopops) would you drink?" | 4462 |
| Mental health conditions ever diagnosed by a professional | 408 | "Have you been diagnosed with one or more of the following mental health conditions by a professional, even if you don't have it currently? By professional we mean any doctor, nurse or person with specialist training (such as a psychologist or therapist). Please include conditions even if you did not need treatment for them or if you did not agree with the diagnosis. Select all that apply." Participants were offered 3 sets of options which have been combined here into a single answer. The following conditions were in group A: Depression; Mania, hypomania, bipolar or manic-depression; Schizophrenia; Any other type of psychosis or psychotic illness; A personality disorder; Autism, Asperger's or autistic spectrum disorder; Attention deficit or attention deficit and hyperactivity disorder; Obsessive compulsive disorder (OCD) The following conditions were in group B: Anxiety or nerves; Generalized anxiety disorder; Social anxiety or social phobia; Agoraphobia; Any other phobia (e.g. disabling fear of heights or spiders); Panic attacks; Panic disorder; Post traumatic stress disorder (PTSD) The following conditions were in group C: Anorexia nervosa; Bulimia nervosa; Binge-eating disorder; Any other eating disorder” | 29000 |
| Mental health conditions ever experienced by first degree blood relatives | 5,332 | "To your knowledge, have any of your first-degree blood relatives had any of the following conditions? [Participant was offered a set of options which have been combined here into a single answer.] Select all that apply." | 29001 |

*Overall monthly alcohol intake was derived by summing these individual measures. Measure details are reflected from the UK Biobank Showcase database.^1^

**UVMR Results**

*Supplementary Table S3*. F statistics from bidirectional UVMR analyses of autism, attention deficit hyperactivity disorder (ADHD) and depression on alcohol consumption

| **Exposure** | **Outcome** | **NSNPs** | **F statistic** |
| --- | --- | --- | --- |
| Autism | Alcohol consumption | 5 | 28.10 |
| ADHD | Alcohol consumption | 21 | 36.60 |
| Depression | Alcohol consumption | 89 | 40.40 |
| Alcohol consumption | Autism | 10 | 24.70 |
| Alcohol consumption | ADHD | 10 | 24.70 |
| Alcohol consumption | Depression | 10 | 24.70 |

*Supplementary Table S4*. Heterogeneity statistics from bidirectional UVMR analyses of autism, attention deficit hyperactivity disorder (ADHD) and depression on alcohol consumption

| **Exposure** | **Outcome** | **IVW-MR** | | | **MR-Egger** | | |
| --- | --- | --- | --- | --- | --- | --- | --- |
|  |  | Q | Q df | P-value | Q | Q df | P-value |
| Autism | Alcohol consumption | 1.93 | 4 | 0.748 | 1.91 | 3 | 0.591 |
| ADHD | Alcohol consumption | 31.19 | 20 | 0.053 | 30.60 | 19 | 0.045 |
| Depression | Alcohol consumption | 120.79 | 88 | 0.011 | 120.66 | 87 | 0.010 |
| Alcohol consumption | Autism | 15.90 | 9 | 0.070 | 10.50 | 8 | 0.234 |
| Alcohol consumption | ADHD | 22.70 | 9 | 0.007 | 18.20 | 8 | 0.020 |
| Alcohol consumption | Depression | 25.70 | 9 | 0.002 | 25.40 | 8 | 0.001 |

*Supplementary Table S5*. MR-Egger intercept test for pleiotropic effects from bidirectional UVMR analyses of autism, attention deficit hyperactivity disorder (ADHD) and depression on alcohol consumption

| **Exposure** | **Outcome** | **MR-Egger intercept** | **P value** |
| --- | --- | --- | --- |
| Autism | Alcohol consumption | 0.002 | 0.893 |
| ADHD | Alcohol consumption | 0.004 | 0.553 |
| Depression | Alcohol consumption | -0.001 | 0.756 |
| Alcohol consumption | Autism | 0.073 | 0.077 |
| Alcohol consumption | ADHD | 0.014 | 0.196 |
| Alcohol consumption | Depression | 0.004 | 0.749 |

*Supplementary Table S6*. Results from bidirectional UVMR analyses of autism, attention deficit hyperactivity disorder (ADHD) and depression on alcohol consumption after Steiger filtering (IVW-MR and MR-Egger)

| **Exposure** | **Outcome** | **IVW-MR** | | | **MR-Egger** | | | | | | |
| --- | --- | --- | --- | --- | --- | --- | --- | --- | --- | --- | --- |
|  |  | Effect estimate | CI95% (lower, upper) | P value | | Effect estimate | | | CI95% (lower, upper) | | P value |
| Autism^1^ | Alcohol use | 0.02 | -0.01, 0.05 | 0.125 | |  | -0.00 | -0.32, 0.32 | | 0.998 | |
| ADHD^2^ | Alcohol use | -0.01 | -0.08, 0.07 | 0.866 | |  | -0.21 | -0.86, 0.44 | | 0.543 | |
| Depression^3^ | Alcohol use | -0.01 | -0.04, 0.03 | 0.727 | |  | 0.18 | -0.02, 0.40 | | 0.080 | |
| Alcohol use^4^ | Autism | -0.04 | -1.33, 0.43 | 0.318 | |  | -5.12 | -9.69, -0.55 | | 0.060 | |
| Alcohol use^5^ | ADHD | -0.02 | -0.24, 0.21 | 0.892 | |  | -0.91 | -2.16, 0.35 | | 0.195 | |
| Alcohol use^6^ | Depression | -0.00 | -0.23, 0.22 | 0.978 | |  | -0.24 | -1.66, 1.18 | | 0.749 | |

^1^nSNPs=5, ^2^nSNPs=21, ^3^nSNPs=64, ^4^nSNPs=10, ^5^nSNPs=10, ^6^nSNPs=10

*Supplementary Table S7*. Results from bidirectional UVMR analyses of autism, attention deficit hyperactivity disorder (ADHD) and depression on alcohol consumption after Steiger filtering (Weighted median and Weighted mode)

| **Exposure** | **Outcome** | **Weighted median** | | | | **Weighted mode** | | |
| --- | --- | --- | --- | --- | --- | --- | --- | --- |
|  |  | Effect estimate | CI95% (lower, upper) | P value |  | Effect estimate | CI95% (lower, upper) | P value |
| Autism^1^ | Alcohol use | 0.02 | -0.02, 0.06 | 0.349 |  | 0.02 | -0.04, 0.08 | 0.610 |
| ADHD^2^ | Alcohol use | -0.00 | -0.09, 0.08 | 0.942 |  | -0.00 | -0.17, 0.17 | 0.982 |
| Depression^3^ | Alcohol use | -0.01 | -0.06, 0.04 | 0.614 |  | -0.03 | -0.18, 0.11 | 0.652 |
| Alcohol use^4^ | Autism | -0.71 | -1.60, 0.17 | 0.115 |  | -0.90 | -3.10, 1.27 | 0.437 |
| Alcohol use^5^ | ADHD | -0.10 | -0.27, 0.13 | 0.508 |  | -0.08 | -0.64, 0.47 | 0.774 |
| Alcohol use^6^ | Depression | 0.01 | -0.20, 0.22 | 0.895 |  | 0.12 | -0.54, 0.76 | 0.737 |

^1^nSNPs=5, ^2^nSNPs=21, ^3^nSNPs=64, ^4^nSNPs=10, ^5^nSNPs=10, ^6^nSNPs=10

*Supplementary Table S8*. F statistics from bidirectional UVMR analyses of autism, attention deficit hyperactivity disorder (ADHD) and depression on alcohol consumption after Steiger filtering

| **Exposure** | **Outcome** | **NSNPs** | **F statistic** |
| --- | --- | --- | --- |
| Autism | Alcohol consumption | 5 | 28.10 |
| ADHD | Alcohol consumption | 21 | 36.60 |
| Depression | Alcohol consumption | 64 | 42.00 |
| Alcohol consumption | Autism | 10 | 24.70 |
| Alcohol consumption | ADHD | 10 | 24.70 |
| Alcohol consumption | Depression | 10 | 24.70 |

*Supplementary Table S9*. Heterogeneity statistics from bidirectional UVMR analyses of autism, attention deficit hyperactivity disorder (ADHD) and depression on alcohol consumption after Steiger filtering

| **Exposure** | **Outcome** | **IVW-MR** | | | **MR-Egger** | | |
| --- | --- | --- | --- | --- | --- | --- | --- |
|  |  | Q | Q df | P-value | Q | Q df | P-value |
| Autism | Alcohol consumption | 1.93 | 4 | 0.591 | 1.91 | 3 | 0.748 |
| ADHD | Alcohol consumption | 31.19 | 20 | 0.053 | 30.60 | 19 | 0.044 |
| Depression | Alcohol consumption | 33.75 | 63 | 0.999 | 30.28 | 62 | 0.999 |
| Alcohol consumption | Autism | 15.90 | 9 | 0.070 | 10.50 | 8 | 0.234 |
| Alcohol consumption | ADHD | 22.70 | 9 | 0.007 | 18.20 | 8 | 0.020 |
| Alcohol consumption | Depression | 25.70 | 9 | 0.002 | 25.40 | 8 | 0.001 |

*Supplementary Table S10*. MR-Egger intercept test for pleiotropic effects from bidirectional UVMR analyses of autism, attention deficit hyperactivity disorder (ADHD) and depression on alcohol consumption after Steiger filtering

| **Exposure** | **Outcome** | **MR-Egger intercept** | **P value** |
| --- | --- | --- | --- |
| Autism | Alcohol consumption | 0.002 | 0.892 |
| ADHD | Alcohol consumption | 0.004 | 0.553 |
| Depression | Alcohol consumption | -0.004 | 0.067 |
| Alcohol consumption | Autism | 0.073 | 0.077 |
| Alcohol consumption | ADHD | 0.014 | 0.196 |
| Alcohol consumption | Depression | 0.004 | 0.749 |

*Supplementary Table S11*. Results from bidirectional UVMR analyses of autism and depression on alcohol consumption after removing SNPs rs112635299 and rs35277013 (IVW-MR pre and post Steiger filtering)

| **Exposure** | **Effect estimate (beta)** | **CI95% (lower, upper)** | **P value** |
| --- | --- | --- | --- |
| Autism^1^ | 0.02 | -0.01, 0.05 | 0.125 |
| Depression^1^ | 0.02 | -0.02, 0.05 | 0.348 |
| Autism^2^ | 0.02 | -0.01, 0.05 | 0.125 |
| Depression^2^ | -0.01 | -0.04, 0.03 | 0.727 |

^1^Pre Steiger filtering, ^2^ Post Steiger filtering

*Supplementary Table S12*. Results from bidirectional UVMR analyses of autism and depression on alcohol consumption after removing SNPs rs112635299 and rs35277013 (MR-Egger pre and post Steiger filtering)

| **Exposure** | **Effect estimate (beta)** | **CI95% (lower, upper)** | **P value** |
| --- | --- | --- | --- |
| Autism^1^ | -0.00 | -0.32, 0.32 | 0.998 |
| Depression^1^ | 0.05 | -0.15, 0.25 | 0.643 |
| Autism^2^ | -0.00 | -0.32, 0.32 | 0.998 |
| Depression^2^ | 0.18 | -0.02, 0.39 | 0.080 |

^1^Pre Steiger filtering, ^2^ Post Steiger filtering

*Supplementary Table S13*. Results from bidirectional UVMR analyses of autism and depression on alcohol consumption after removing SNPs rs112635299 and rs35277013 (Weighted median pre and post Steiger filtering)

| **Exposure** | **Effect estimate (beta)** | **CI95% (lower, upper)** | **P value** |
| --- | --- | --- | --- |
| Autism^1^ | 0.02 | -0.02, 0.06 | 0.356 |
| Depression^1^ | -0.01 | -0.05, 0.04 | 0.746 |
| Autism^2^ | 0.02 | -0.02, 0.06 | 0.349 |
| Depression^2^ | -0.01 | -0.06, 0.03 | 0.609 |

^1^Pre Steiger filtering, ^2^ Post Steiger filtering

*Supplementary Table S14*. Results from bidirectional UVMR analyses of autism and depression on alcohol consumption after removing SNPs rs112635299 and rs35277013 (Weighted mode pre and post Steiger filtering)

| **Exposure** | **Effect estimate (beta)** | **CI95% (lower, upper)** | **P value** |
| --- | --- | --- | --- |
| Autism^1^ | 0.02 | -0.04, 0.08 | 0.616 |
| Depression^1^ | -0.03 | -0.16, 0.11 | 0.684 |
| Autism^2^ | 0.02 | -0.04, 0.08 | 0.610 |
| Depression^2^ | -0.03 | -0.19, 0.12 | 0.661 |

^1^Pre Steiger filtering, ^2^ Post Steiger filtering

**MVMR Results**

*Supplementary Table S15*. Conditional F statistics from multivariable MR analyses of autism, attention deficit hyperactivity disorder (ADHD) and depression on alcohol consumption

| **Exposure** | **Outcome** | **F statistic** |
| --- | --- | --- |
| Autism | Alcohol consumption | 2.83 |
| ADHD | Alcohol consumption | 6.65 |
| Depression | Alcohol consumption | 10.66 |

*Supplementary Table S16*. Cochran’s Q statistics from multivariable MR analyses of autism, attention deficit hyperactivity disorder (ADHD) and depression on alcohol consumption

| **Q** | **Qpval** |
| --- | --- |
| 115 | 0.050 |

*Supplementary Table S17*. Qhet estimates from multivariable MR analyses of autism, attention deficit hyperactivity disorder (ADHD) and depression on alcohol consumption

| **Exposure** | **Estimates** |
| --- | --- |
| Autism | 0.01 |
| ADHD | 0.08 |
| Depression | -0.03 |

*Supplementary Table S18*. Results from multivariable MR-Egger analyses of autism, attention deficit hyperactivity disorder (ADHD) and depression on alcohol consumption

| **Exposure** | **Effect estimate (beta)** | **CI95% (lower, upper)** | **P value** |
| --- | --- | --- | --- |
| Autism | -0.01 | -0.06, 0.03 | 0.375 |
| ADHD | -0.03 | -0.05, 0.12 | 0.310 |
| Depression | 0.01 | -0.02, 0.04 | 0.529 |

*Supplementary Table S19*. Results from multivariable IVW-MR analyses of autism, attention deficit hyperactivity disorder (ADHD) and depression on alcohol consumption after Steiger filtering

| **Exposure** | **Effect estimate (beta)** | **CI95% (lower, upper)** | **P value** |
| --- | --- | --- | --- |
| Autism | 0.01 | -0.01, 0.03 | 0.538 |
| ADHD | 0.03 | -0.03, 0.10 | 0.289 |
| Depression | -0.01 | -0.03, 0.03 | 0.361 |

*Supplementary Table S20*. Conditional F statistics from multivariable MR analyses of autism, attention deficit hyperactivity disorder (ADHD) and depression on alcohol consumption after Steiger filtering

| **Exposure** | **Outcome** | **F statistic** |
| --- | --- | --- |
| Autism | Alcohol consumption | 3.12 |
| ADHD | Alcohol consumption | 7.36 |
| Depression | Alcohol consumption | 12.85 |

*Supplementary Table S21*. Cochran’s Q statistics from multivariable MR analyses of autism, attention deficit hyperactivity disorder (ADHD) and depression on alcohol consumption after Steiger filtering

| **Q** | **Qpval** |
| --- | --- |
| 34.40 | 1.00 |

*Supplementary Table S22*. Qhet estimates from multivariable MR analyses of autism, attention deficit hyperactivity disorder (ADHD) and depression on alcohol consumption after Steiger filtering

| **Exposure** | **Estimates** |
| --- | --- |
| Autism | -0.01 |
| ADHD | 0.04 |
| Depression | 0.00 |

*Supplementary Table S23*. Results from multivariable MR-Egger analyses of autism, attention deficit hyperactivity disorder (ADHD) and depression on alcohol consumption after Steiger filtering

| **Exposure** | **Effect estimate (beta)** | **CI95% (lower, upper)** | **P value** |
| --- | --- | --- | --- |
| Autism | -0.01 | -0.06, 0.03 | 0.375 |
| ADHD | 0.03 | -0.05, 0.12 | 0.310 |
| Depression | 0.01 | -0.02, 0.04 | 0.529 |

*Supplementary Table S24*. Conditional F statistics of genetic liability for autism and ADHD in pairwise multivariable MR analyses

| **Exposure** | **Outcome** | **F statistic** |
| --- | --- | --- |
| Autism | Alcohol consumption | 5.76 |
| ADHD | Alcohol consumption | 12.86 |

*Supplementary Table S25*. Conditional F statistics of genetic liability for autism and depression in pairwise multivariable MR analyses

| **Exposure** | **Outcome** | **F statistic** |
| --- | --- | --- |
| Autism | Alcohol consumption | 2.69 |
| Depression | Alcohol consumption | 10.20 |

*Supplementary Table S26*. Conditional F statistics of genetic liability for ADHD and depression in pairwise multivariable MR analyses

| **Exposure** | **Outcome** | **F statistic** |
| --- | --- | --- |
| ADHD | Alcohol consumption | 7.71 |
| Depression | Alcohol consumption | 16.45 |

*Supplementary Table S27*. Results from multivariable IVW-MR analyses of autism and attention deficit hyperactivity disorder (ADHD) on alcohol consumption

| **Exposure** | **Effect estimate (beta)** | **CI95% (lower, upper)** | **P value** |
| --- | --- | --- | --- |
| Autism^1^ | 0.01 | -0.03, 0.05 | 0.670 |
| ADHD^2^ | -0.01 | -0.11, 0.09 | 0.845 |

^1^nSNPs=2, ^2^nSNPs=21

*Supplementary Table S28.* Results from multivariable IVW-MR analyses of autism and attention deficit hyperactivity disorder (ADHD) on alcohol consumption after Steiger filtering

| **Exposure** | **Effect estimate (beta)** | **CI95% (lower, upper)** | **P value** |
| --- | --- | --- | --- |
| Autism^1^ | 0.01 | -0.03, 0.05 | 0.670 |
| ADHD^2^ | -0.01 | -0.11, 0.09 | 0.845 |

^1^nSNPs=2, ^2^nSNPs=21

*Supplementary Table S29*. Results from multivariable IVW-MR analyses of autism and depression on alcohol consumption

| **Exposure** | **Effect estimate (beta)** | **CI95% (lower, upper)** | **P value** |
| --- | --- | --- | --- |
| Autism^1^ | 0.02 | -0.01, 0.06 | 0.473 |
| Depression^2^ | -0.01 | -0.04, 0.02 | 0.224 |

^1^nSNPs=2, ^2^nSNPs=89

*Supplementary Table S30*. Results from multivariable IVW-MR analyses of autism and depression on alcohol consumption after Steiger filtering

| **Exposure** | **Effect estimate (beta)** | **CI95% (lower, upper)** | **P value** |
| --- | --- | --- | --- |
| Autism^1^ | -0.00 | -0.01, 0.04 | 0.155 |
| Depression^2^ | -0.01 | -0.04, 0.01 | 0.313 |

^1^nSNPs=2, ^2^nSNPs=64

*Supplementary Table S31.* Results from multivariable IVW-MR analyses of attention deficit hyperactivity disorder (ADHD) and depression on alcohol consumption

| **Exposure** | **Effect estimate (beta)** | **CI95% (lower, upper)** | **P value** |
| --- | --- | --- | --- |
| ADHD^1^ | 0.02 | -0.06, 0.01 | 0.625 |
| Depression^2^ | 0.01 | -0.03, 0.06 | 0.524 |

^1^nSNPs=11, ^2^nSNPs=86

*Supplementary Table S32*. Results from multivariable IVW-MR analyses of attention deficit hyperactivity disorder (ADHD) and depression on alcohol consumption after Steiger filtering

| **Exposure** | **Effect estimate (beta)** | **CI95% (lower, upper)** | **P value** |
| --- | --- | --- | --- |
| ADHD^1^ | 0.03 | -0.02, 0.09 | 0.267 |
| Depression^2^ | -0.01 | -0.04, 0.02 | 0.517 |

^1^nSNPs=10, ^2^nSNPs=61

**References**

1. Showcase Homepage. Accessed August 25, 2026. https://biobank.ndph.ox.ac.uk/ukb/index.cgi
